# Normalisation of SARS-CoV-2 concentrations in wastewater: the use of flow, conductivity and CrAssphage

**DOI:** 10.1101/2021.11.30.21266889

**Authors:** Jeroen Langeveld, Remy Schilperoort, Leo Heijnen, Goffe Elsinga, Claudia E.M. Schapendonk, Ewout Fanoy, Evelien I.T. de Schepper, Marion P. G. Koopmans, Miranda de Graaf, Gertjan Medema

**Author notes:** Corresponding author: Jeroen Langeveld.

## Abstract

Over the course of the COVID-19 pandemic in 2020-2021, monitoring of SARS-CoV-2 RNA in wastewater has rapidly evolved into a supplementary surveillance instrument for public health. Short term trends (2 weeks) are used as a basis for policy and decision making on measures for dealing with the pandemic. Normalization is required to account for the varying dilution rates of the domestic wastewater, that contains the shedded virus RNA. The dilution rate varies due to runoff, industrial discharges and extraneous waters. Three normalization methods using flow, conductivity and CrAssphage, have been investigated on 9 monitoring locations between Sep 2020 and Aug 2021, rendering 1071 24-hour flow-proportional samples. In addition, 221 stool samples have been analyzed to determine the daily CrAssphage load per person. Results show that flow normalization supported by a quality check using conductivity monitoring is the advocated normalization method in case flow monitoring is or can be made available. Although Crassphage shedding rates per person vary greatly, the CrAssphage loads were very consistent over time and space and direct CrAssphage based normalization can be applied reliably for populations of 5600 and above.

## 1. Introduction

Over the course of the COVID-19 pandemic in 2020-2021, monitoring of SARS-CoV-2 RNA in wastewater has rapidly evolved into a supplementary surveillance instrument for public health. (Lodder and de Roda Husman 2020, Medema et al. 2020b, Kirby et al., 2021). It is currently used in many countries (<u>COVIDPoops19 Dashboard | covid19wbec.org</u>) at different scales, from national surveillance at all (<u>Virus particles in wastewater | Coronavirus Dashboard | Government.nl</u>) or selected wastewater treatment plants (<u>Szennyvizekben mért SARS-CoV-2 vírus koncentrációja (gov.hu)</u>; <u>Koronaviruksen jätevesiseurannan viikkoraportti (thl.fi)</u>; <u>Données ouvertes - Réseau OBEPINE (reseau-obepine.fr)</u>; <u>Coronastep | Luxembourg Institute of Science and Technology (list.lu)</u>; <u>SARS-CoV-2 in Wastewater (sensors-eawag.ch)</u>) to regional (Wolfe et al., 2021; Ai et al., 2021;<u>Sarsaigua (icra.cat)</u>), (sub) city level (Rasero et al., 2021; Yaniv et al., 2021) and building level (Davo et al., 2021; Sweetapple et al., 2021; Betancourt et al., 2021). The concentration of SARS-CoV-2 RNA in wastewater has shown to reflect and even precede the trends of the newly reported cases or COVID-19 hospitalizations (Medema et al. 2020a; Prado, Fumian et al. 2021; Ho et al., 2021). In situations with low COVID-19 prevalence, wastewater surveillance is being used as an early warning system (Betancourt et al. 2020, Medema et al. 2020a, Ahmed et al. 2021) enabling rapid and targeted measures to limit the COVID-19 transmission. Wastewater surveillance is also an efficient tool to monitor the emergence of (signature mutations of) new variants-of-concern in communities, using targeted sequencing (Rios et al., 2021; Rothman et al., 2021; Jahn et al., 2021) or targeted PCR methods (Graber et al., 2021, Heijnen et al. 2021). A key advantage of wastewater surveillance is the possibility to obtain objective information about virus circulation in a community. The ‘traditional’ surveillance of reported cases is subject to bias by (changes in) testing strategies, access to testing, and compliance of communities, and reflect only those that get tested. SARS-CoV-2 concentrations in community wastewater are independent of testing behavior, thereby providing a more complete and objective image of the virus circulation in the community provided there is unrestricted access to sanitation.

Public health agencies use the wastewater signal to spot changes in testing behavior over time or between communities (de Graaf et al, in preparation) and to verify the trends in reported cases in communities. Decision-making on (partial) lockdowns and travel restrictions requires up-to-date and reliable information on trends in virus circulation, preferably at a high spatial resolution. The relevant time window of many of these decisions is the information of 1 -2 antecedent weeks; i.e. the weekly updates of the epidemiological situation (e.g. https://www.rivm.nl/en/coronavirus-covid-19/weekly-figures) and the maps in support of the Council Recommendation on a coordinated approach to travel measures in the EU at https://www.ecdc.europa.eu/en/covid-19/situation-updates/weekly-maps-coordinated-restriction-free-movement, that is based on the testing, positivity and notification rate over a time frame of 2 weeks. To monitor trends in SARS-CoV-2 circulation via wastewater in a similar time frame, frequent and representative sampling of a community is necessary. Sample collection typically takes place as 24 h composite samples to account for the typical diurnal pattern associated with wastewater production and toilet use (Medema et al., 2020b; Ort et al. 2010). In addition, wastewater concentrations can be affected by the dilution of domestic wastewater in other water in the sewerage network, such as stormwater runoff, groundwater or industrial discharges. This dilution may vary in time; wet weather discharges can be many times the dry weather discharge of sewers. To account for this variable dilution, normalisation of the measured concentrations by the (daily) flow of wastewater into daily loads is a common procedure in Wastewater Based Epidemiology (WBE), building on methods and procedures developed over the last decade mainly for drug use monitoring (Castiglioni et al., 2014). For SARS-CoV-2 normalisation, the measured RNA concentration in the 24h composite sample can be multiplied by the wastewater flow of the last 24h at the sampling site to compute the daily viral load. This load can in turn be normalised for the population of the sewershed to be able to compare the loads between different areas. The underlying assumption of this flow normalisation approach (Castiglioni et al., 2014) is that the sample taken is representative of the domestic wastewater volume being discharged by the population served by the pumping station or wastewater treatment plant (WWTP). However, there are several situations where this assumption does not hold. They may be attributed to short term changes in population size due to commuters or tourism, or to sewer system dynamics, such as spills of SSOs (sanitary sewer overflows) due to blockages, spills of CSOs during heavy rainfall, transport delays in pressure mains which vary strongly during dry weather and wet weather or delays in the transport of wastewater to the WWTP due to pump failures or other operational issues. In these situations, the wastewater flow at the inlet of the wastewater treatment plant is not representing the amount of wastewater that is produced in that sewershed on that day. Consequently, there is a need for additional quality control in WBE. Normalisation by two independent parameters could be the way forward to check for representativeness of the sample for the shedding of SARS-CoV-2. In addition, in many locations flow data is unavailable, resulting in a need for other types of normalisation parameters.

Many examples of parameters that may be used to normalise wastewater samples are available. Launay et al. (2016) propose the use of electric conductivity (EC) as a proxy of the dilution of wastewater due to storm water or ground water dilution. They showed that the dilution calculated based on EC gives similar results as the use of inert human wastewater tracers such as ibuprofen, naproxen and diclofenac. Many other human wastewater tracers are less applicable as tracer due to e.g. a higher back ground variation or degradation in the sewer (Gao et al., 2017). EC was selected as a cheap parameter which can be reliably measured. Tandukar et al. (2020), Bivins et al. (2020) and Medema et al. (2020b) assessed the applicability of several chemical and biological markers for human input in wastewater.

Like others (Crank et al., 2020; Hillary et al., 2021; Wilder et al., 2021; Heijnen et al., 2021), we selected CrAssphage, given it is a highly abundant virus (almost) exclusively found in human feces and is present in humans world-wide (Edwards et al., 2019). High CrAssphage concentrations are reported in domestic wastewater globally, and the concentrations do not show significant seasonal variation (Balleste et al., 2019), making them a potentially useful index for the human fecal fraction of wastewater.

The aim of this study was to develop a procedure for quality control and normalisation of wastewater samples, in order to allow quantitative trend analysis of SARS-CoV-2 in support of public health decision-making. We compared and evaluated three methods for normalisation: the ‘standard’ daily wastewater flow rate and population normalisation, electric conductivity (EC) as a proxy for dilution and the human fecal biomarker crAssphage. In addition, we analysed whether a combination of two normalisation methods can be used as a quality check and normaliser of SARS-CoV-2 RNA concentration data.

## 2. Material and methods

### 2.1. Wastewater surveillance Rotterdam Rijnmond

In and around the city of Rotterdam (the Netherlands), 9 catchment areas were selected for wastewater sampling (Figure 1). Four catchment areas were sampled at the wastewater treatment plant Dokhaven (INF2, INF3, INF4 and INF5-6), sampling from the pressure mains arriving at the Wastewater Treatment Plants (WWTP). These four areas make up about half of the urban area of the city of Rotterdam and vary in population between 27.044 and 138.280 people (CBS, 2020). Catchment area Pretorialaan (71.325 inhabitants) constitutes roughly half of the Dokhaven INF3 area, and catchment area Katendrecht (4.884 inhabitants) is a small subcatchment of the Pretorialaan catchment. Both catchments are sampled at their respective sewer pumping stations serving the area. The sewer system is cascading meaning that wastewater from Katendrecht is first sampled at sewer pumping station Katendrecht, then again at sewer pumping station Pretorialaan (diluted with wastewater from the rest of the Pretorialaan area), and finally again at Dokhaven INF3 (further diluted). Outside the WWTP Dokhaven area, three predominantly residential areas have been selected for surveillance: Ommoord (28.434 inhabitants), Bergschenhoek (18.750 inhabitants) and Rozenburg (12.374 inhabitants). Ommoord and Bergschenhoek were sampled at their respective sewer pumping stations, Rozenburg at the WWTP serving exclusively the Rozenburg area.

**Figure 1.**
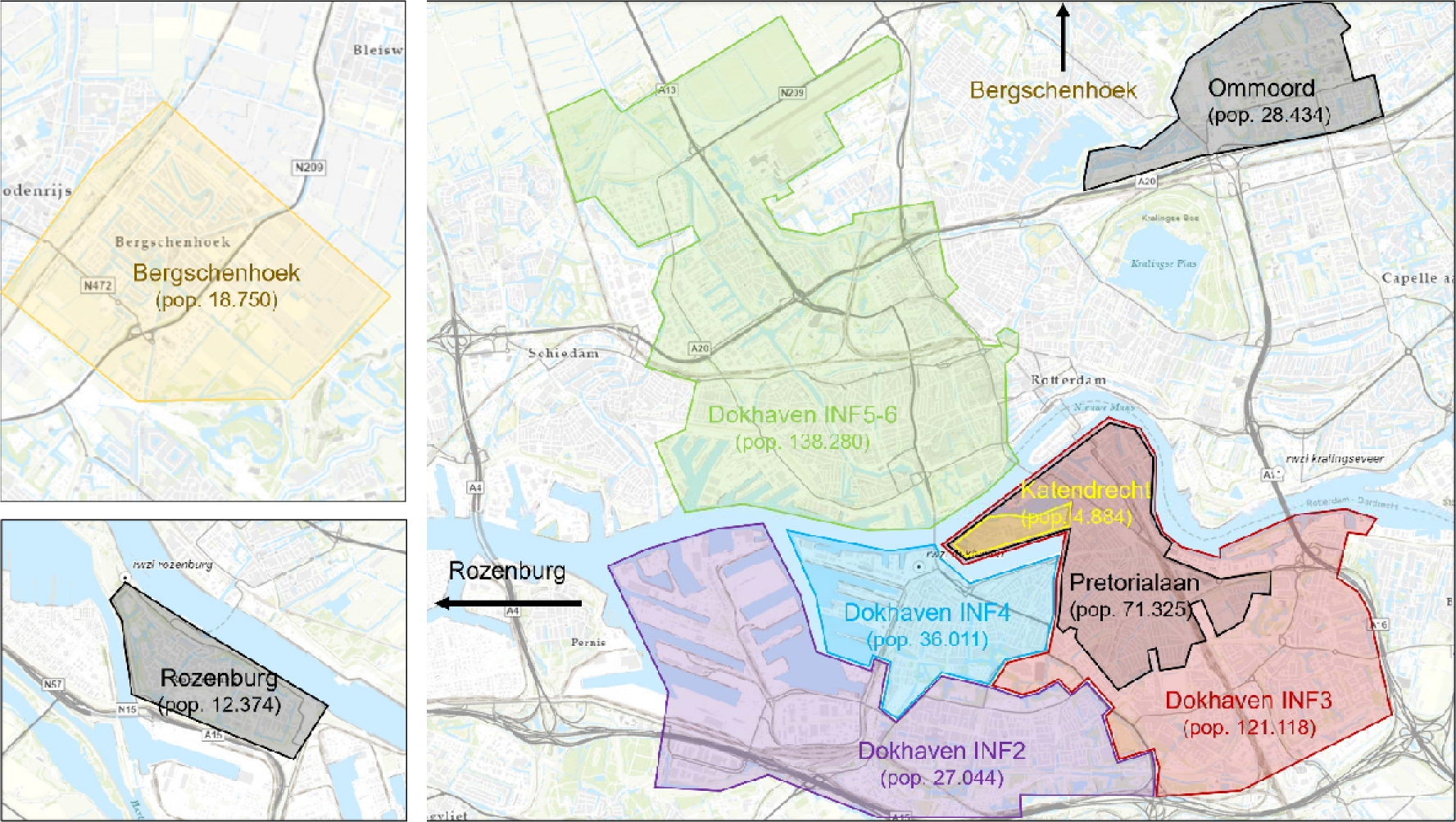
Catchment areas in the Rotterdam Rijnmond area used for wastewater surveillance with their population size.

The sewer network in the Rotterdam area is largely combined, with separate sewer only in more recently developed and renovated areas.

### 2.2. Wastewater sampling and data

At all sampling sites automated composite samplers (Endress+Hauser Liquistation CSF48 and ASP2000 stations) were used to collect flow-proportional 24h composite wastewater samples. The autosamplers were programmed to collect 50 mL aliquots per fixed volume of wastewater (as measured by the flow sensors at the WWTPs/pumping stations). This fixed volume varied between 12 m^3^ (Katendrecht) and 170 m^3^ (Dokhaven INF3) and was selected to ensure a minimum of 100 aliquots (5 L of sample) for each 24h sample throughout the year (NEN6600). All locations followed the same sampling schedule: three 24h samples per week (Sun 08h00 - Mon 08h00, Tue 08h00 - Wed 08h00 and Thu 08h00 - Fri 08h00) from September 2020 (Bergschenhoek: January 2021) to August 2021. Samples were stored inside the autosampler at a temperature between 1°C and 5°C until sample collection (Mon, Wed and Fri between 08h00 and 16h00). At the time of sample collection, the collected composite sample was mixed by manual stirring and an aliquot of 250 mL was collected from the container in the autosampler, stored in a sterile flask and transported at 4°C to the laboratory.

Upon sample collection the quality of each sample was assessed comparing the theoretical sample volume (number of aliquots in 24h, multiplied by aliquot volume) with the actual sample volume. In case of sampling ‘failure’ (> 7,5% difference according to NEN6600), details of the sampling process were further studied to determine whether the sample could still be included in the data set. Overall approximately 5-7% (depending on the location) of all sampling failed completely (not a single aliquot in the container in a 24h period) due to e.g. a power cut, maintenance activities, clogging of the autosampler, public holidays etc. For another 5-15 % of samples a deviation of more than 7,5% was observed. These were caused mainly by (partial or temporal) clogging of the equipment (relatively often coinciding with storm events) or logistical errors in the field. Overall, 89,6 % of potential samples could be used for processing.

### 2.3. Virus concentration and nucleic acid extraction

Samples were processed within one week after sampling using the procedure as previously described (Medema et al 2020). In short, centrifugation was used as pre-treatment to remove larger particles. Virus particles were concentrated from 50 ml supernatant by ultrafiltration through Centricon® Plus-70 centrifugal ultrafilters with a cut-off of 30 kDa (Millipore, Amsterdam, Netherlands). Mouse Hepatitis Virus (MHV)-A59 (Department Medical Microbiology, Leiden University Medical Center, Leiden, Netherlands) was spiked to each concentrate as quality control. Nucleic acid was extracted from the concentrate with the Biomerieux Nuclisens kit (Biomerieux, Amersfoort, Netherlands) in combination with the semi-automated KingFisher mL (Thermo Scientific, Bleiswijk, Netherlands) as previously described. Extracted nucleic acid was eluted in a volume of 100 µl.

### 2.4. RT-qPCR for Coronavirus RNA-quantification in wastewater

The N2 and E_Sarbecco gene fragments of SARS-CoV-2 were used as qRT-PCR targets, N2 was used for quantification, E_Sarbecco for confirmation (both needed to be present for a positive result). Reagents and reaction conditions were as previously described (Medema et al. 2020; Heijnen et al. 2021). All RT-PCR’s were run as technical duplicates on 5 µl extracted nucleic acid. Reactions were considered positive if the cycle threshold was below 40 cycles. Spiked MHV-A59 RNA was detected by performing a MHV-A59 specific RT-qPCR targeting the N-gene using the primers and reaction profile described by (Raabenet al. 2007).

### 2.5. PCR for CrAssphage quantification in wastewater and feces

A CrAssphage CPQ_064 specific PCR (Stachler, Kelty et al. 2017) was used to quantify this DNA-virus. Assays were performed in duplicate on 5 µl 1:10 diluted extracted nucleic acid as described previously (Heijnen et al. 2021).

### 2.6. PCR for CrAssphage quantification in stool samples

Feces swabs from 221 patients from the Rotterdam Rijnmond region that were tested positive for SARS-CoV-2, were collected in 3 ml of viral transport medium. Samples were spun down for 5 min at 17,000 g, nucleic acids were isolated from 200μl supernatant using the high pure RNA isolation kit (Roche) while omitting the DNAse I step. The RT-PCR described in 2.5 was used to quantify levels of CrAssphage. This study was approved by the Medical Ethical Committee of Erasmus MC under MEC-2020-0617

### 2.7. Normalisation procedure

In the study, three normalisation procedures have been developed to be able to assess flow, EC and crAssphage as normalisation parameters. In each procedure, the percentage of *domestic* wastewater in was determined. The flow based normalisation procedure comprises the following steps:

1. Preparation step: determine the number of inhabitants in the catchment and determine the domestic dry weather flow (ddwf) expressed in litres per person per day. In the Netherlands, the ddwf is very stable at 120 l pppd (www.riool.net). In other countries, the ddwf may vary per location and can typically be obtained via local sewer authorities.
2. Determine the proportion of domestic sewage in a wastewater sample using flow data:

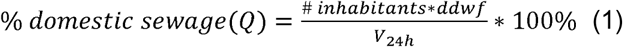

where ddwf = average daily domestic wastewater production per person, V_24h_ = measured wastewater volume over 24 h sampling time

The EC based normalisation procedure requires more additional data to enable direct comparison with the flow base normalisation:

1. Determine number of inhabitants, the average domestic dry weather flow in litres per person per day and the average sewage dry weather flow, which is used as a reference to account for the dilution with extraneous water during dry weather. This average dry weather flow can be extracted as the 40^th^ percentile from a time series of flow data at the sampling site (Mulder et al., 2020). For most monitoring locations, the difference between the 30^th^ and 50^th^ percentile value is relatively small. For catchments with a strong seasonal variation of the amount of extraneous waters, it may be necessary to derive a reference for the wet and dry season. This assessment is easy to make using the methods as described in Weiß et al, 2002.
2. Determine the reference EC value during dry weather flow by taking the average of all days with a daily flow volumes ranging between the 10^th^ and 50^th^ percentile derived from a 1 year time series. Days with flows smaller than the 10^th^ percentile could be affected by operational irregularities, such pump failures or drinking water outage, while flows above the 50^th^ percentile could be affected by (small) storm events.
3. Determine the proportion of domestic sewage in a wastewater sample using EC data:

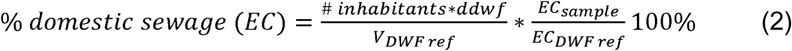

where V_DWF ref_ = average wastewater volume during dry weather flow (40^th^ percentile of daily volumes derived from 1 year time series of flow monitoring, EC_sample_ = measured EC in sample, EC_DWF ref_ = average EC of all samples with flows ranging between 10^th^ and 50^th^ percentile of daily volumes derived from 1 year time series of flow monitoring). Please note that the V_DWF ref_ also contains industrial wastewater and extraneous water. The first term in equation 2 in fact describes the dilution during dry weather flow, the second term in the equation the additional dilution during storm event.

The crAssphage based normalisation follows the same approach as the EC based normalisation and only differs in the parameter used: CrAssphage instead of EC:

1. The concentration during dry weather flow is taken as a reference to enable calculating the proportion of domestic sewage during wet weather using eq 3).

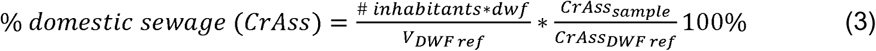

where V_DWF ref_ = average wastewater volume during dry weather flow (40^th^ percentile of daily volumes derived from 1 year time series of flow monitoring), CrAss_sample_ = measured CrAss in sample (genome copies/ml sample), CrAss_DWF ref_ = average CrAss of all samples with flows ranging between 10^th^ and 50^th^ percentile of daily volumes derived from 1 year time series of flow monitoring, (genome copies/ml sample)).

For all three normalisation parameters, normalisation of the SARS-CoV-2 RNA concentration in wastewater is performed by multiplying the measured concentration with the reciprocal of the proportion of domestic sewage in the sample. This is expressed as SARS-CoV-2 N2 RNA concentration (in gene copies) per mL of domestic wastewater, and enables direct comparison of the three normalisation parameters.

## 3. Results and discussion

### 3.1. Normalisation factors differ in time and space

In total, data from 1207 samples from the 9 different city areas from September 2020-August 2021 are available. Flow based normalisation was applied after performing the normalisation procedure by multiplying the lab result (# SARS-CoV-2 N2 gene copies/ml sample) with the reciprocal of the proportion of domestic wastewater. Figure 2 gives an overview of the proportion of domestic wastewater per sample. The large range is due to the fact that all catchments are served by combined sewer systems, which show high dilutions and consequently, low proportions of domestic wastewater, during storm events. As example, Figure 3 shows how storm events affect the observed SARS-CoV-2 concentration for INF3, with a variable normalisation factor up to 2.8. In addition to dilution due to rain water, the median of the correction factors ranges between 2.0 and 3.5. This indicates that also during dry weather a significant proportion of the wastewater sampled consists of non-domestic sources, such as extraneous waters and industrial wastewater. The sewer system of Rotterdam is known for its high proportion of extraneous waters. At WWTP Dokhaven, the average amount of extraneous waters during dry weather flow is 43% of the total influent (Vosse, 2013). Catchment INF 2 Everlo Waalhaven also discharges a substantial amount of industrial wastewater, while catchment Ommoord shows a strongly time-varying amount of extraneous water (see figures S1) with a rather high salinity. This is most likely brackish drainage water in this catchment which is at 5 meters below sea level. The large differences in correction factors implicate that directly comparing measured SARS-CoV-2 concentrations in different city areas without normalisation could result in an incorrect assessment of the relative concentration levels, both within a location over time (trends) and between locations.

**Figure 2.**
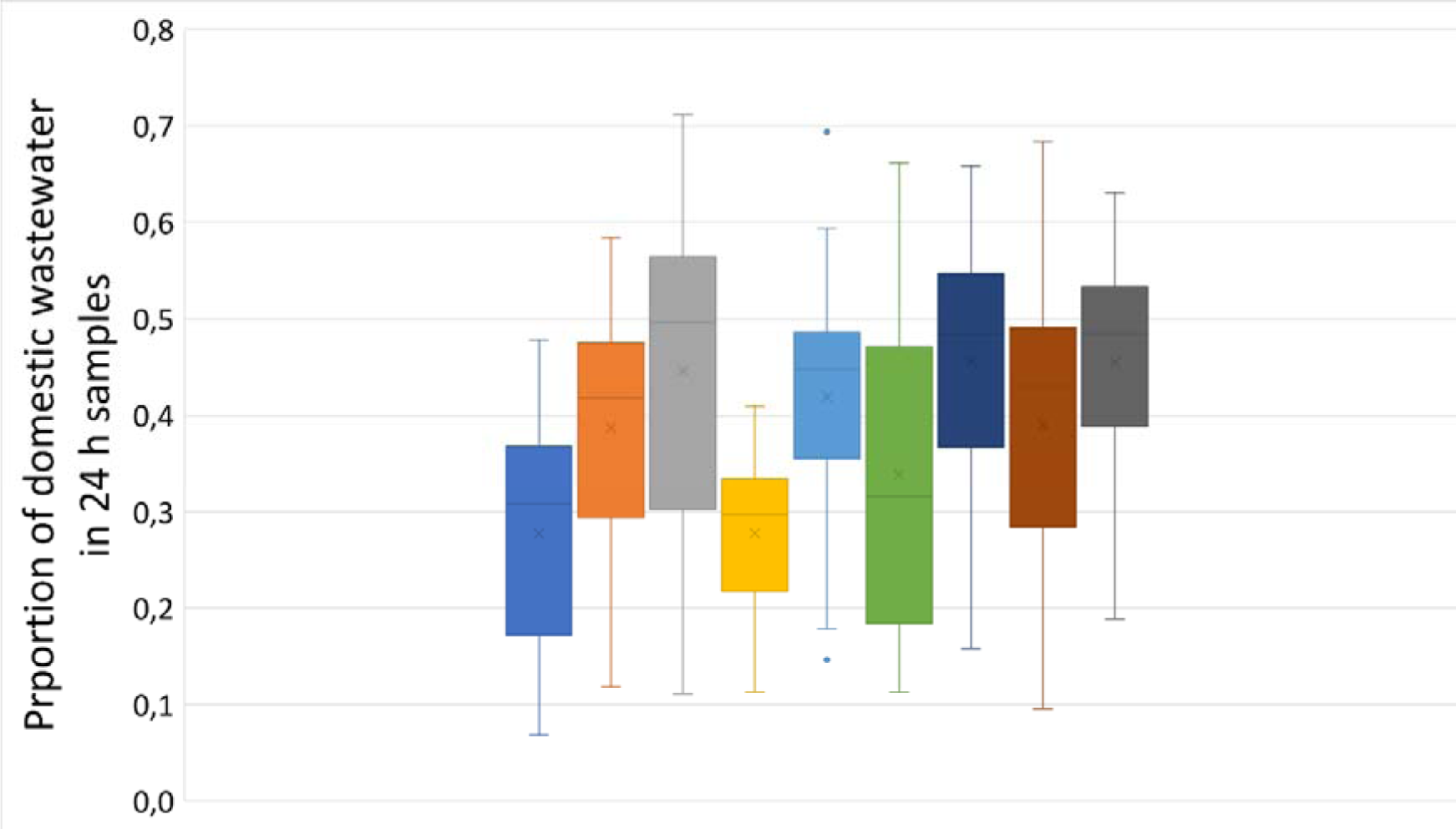
Boxplot with proportion of domestic wastewater in 24h samples. Proportion of domestic wastewater is calculated per sample by dividing the theoretical domestic wastewater production (# inhabitants from figure 1 * 120 lpppd) by the measured 24 flow.

**Figure 3.**
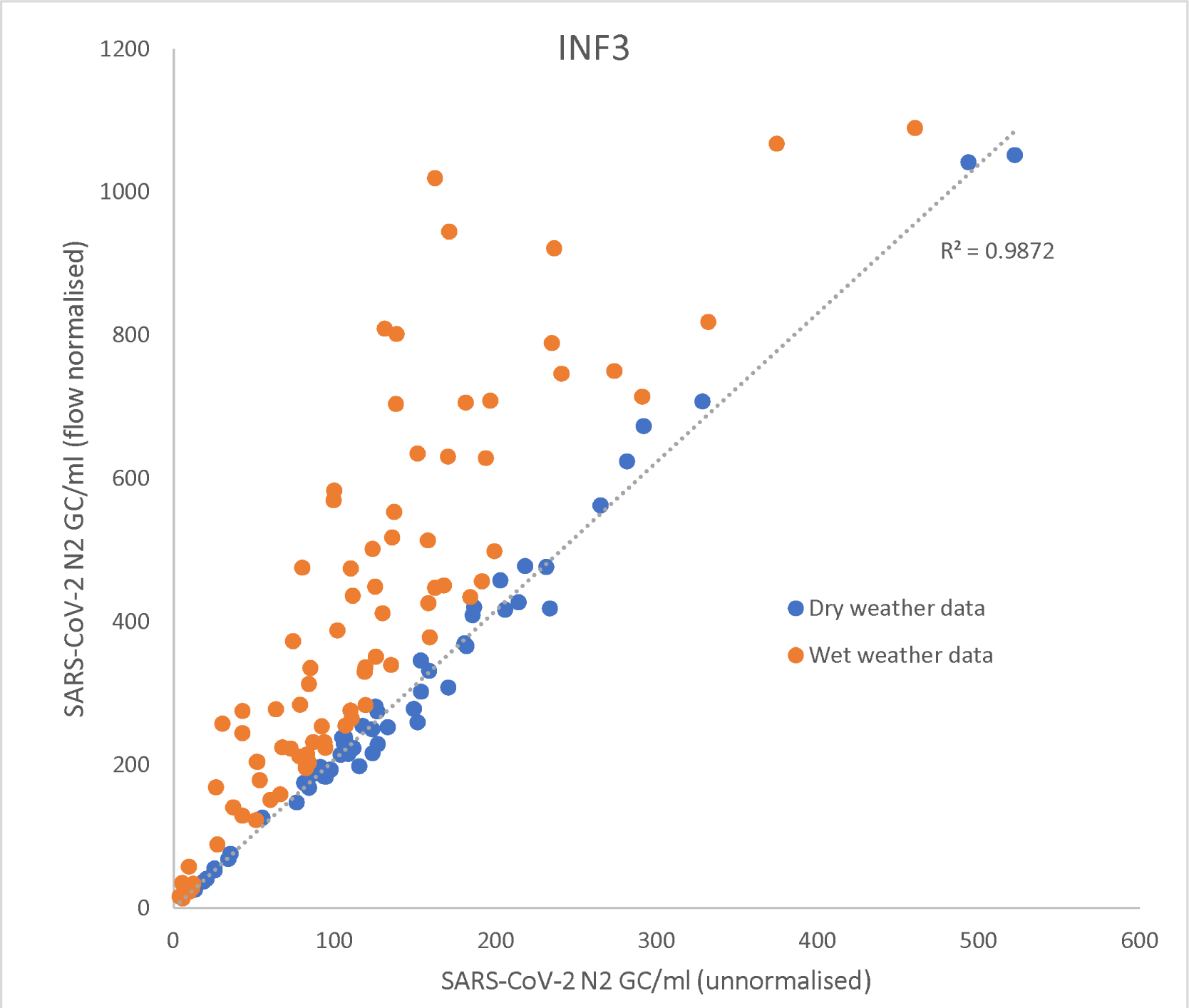
Flow normalised versus unnormalised SARS-CoV-2 concentrations at site INF3 during dry weather and wet weather. The trendline for the dry weather data (flow < V_dwf ref_) shows a high correlation between normalized and unnormalised data, but this is lost in the wet weather data.

### 3.2. Impact of dilution on observed SARS-CoV-2 trends

Flow based normalisation corrects the measured SARS-CoV-2 concentration for the dilution rate of the domestic part of the wastewater. Figure 4 gives an example for the INF3 catchment, all other sites are in the supplementary material. During storm (= high flow) events, the correction factor is higher while on dry days the correction factor is smaller. The long-term trend is not affected, as the catchment in Rotterdam does not show a season-dependent runoff volume and also does not show a seasonal trend in the amount of extraneous water (Vosse, 2013).

**Figure 4.**
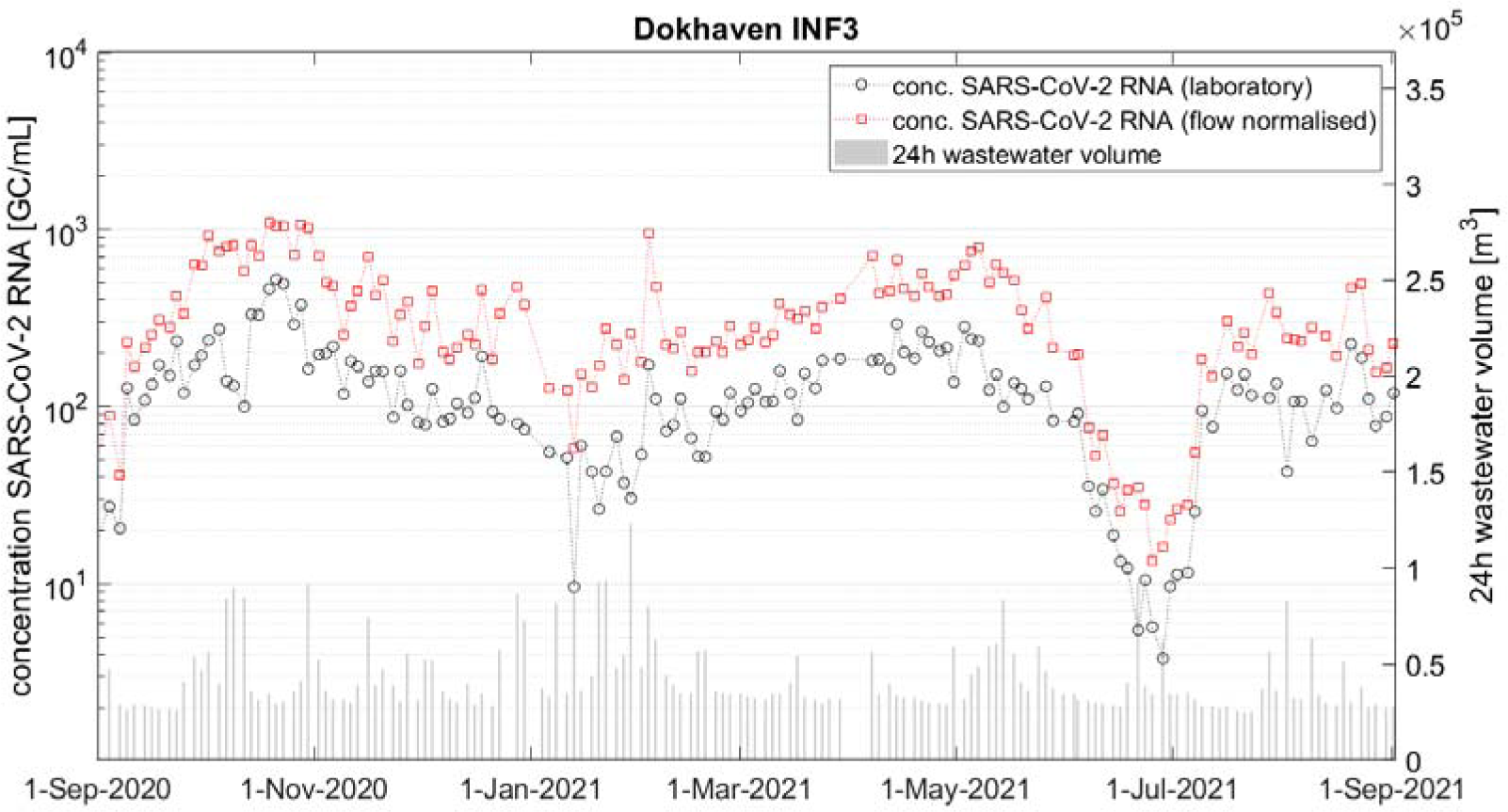
Flow normalised vs unnormalised for catchment INF3

As stated before, the relevant timescale of trend information for public health agencies is in the range of 1-2 weeks. On such a short notice, 2 weeks of consecutive dry weather as well as 2 weeks of consecutive wet weather are both not unlikely. Especially longer periods with wet weather could result in several subsequent samples being diluted. Figure 5 zooms in on the first 4 months of the time series of site INF3. On several occasions, October 5^th^-12^th^, November 11^th^-16^th^, and December 18^th^ –30^th^, uncorrected lab results clearly suggested a downward trend in SARS-CoV-2 levels in the sewage. All were in weeks with rain, and the normalised SARS-CoV-2 levels in domestic sewage in the same periods clearly showed a different trend. From October 30^th^ – November 6^th^, the uncorrected lab results suggested an upward trend, while the normalised SARS-CoV-2 levels showed a downward trend as dilution decreased due to less inflow. Similar differences were observed at the other sites (SI).

**Figure 5.**
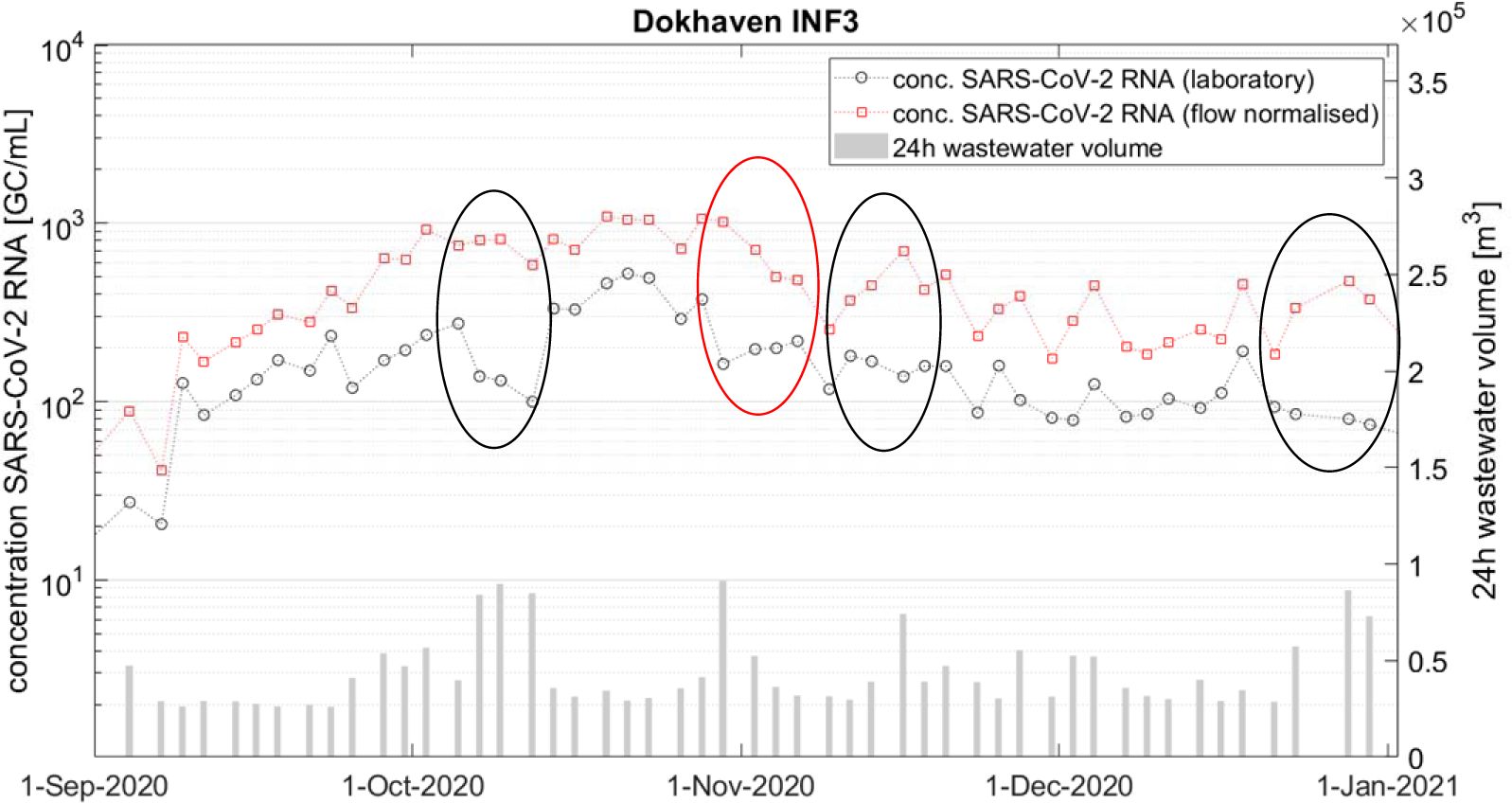
Short-term trends normalised vs unnormalised for catchment INF3. Weeks with a differing trend are indicated

### 3.3. Flow based normalisation versus EC and CrAssphage based normalisation

The three types of normalisation, flow based, EC based and CrAssphage based, result in comparable long term trends in normalised SARS-CoV-2 levels of INF3 (figure 6), apart from a few aberrations. In the second week of February 2021, an increased EC was observed due to road de-icing and as a consequence, the EC based correction factors were lower than the flow based correction factors. CrAssphage based normalisation deviated strongly from flow based and EC based normalisation in the period between March 8^th^ – 15^th^. Figure 7 shows the CrAssphage load per person and day for the entire monitoring period of INF3 (other areas in SI), showing a fairly constant load over the monitoring period, with the exception of the mentioned period. This deviation was observed at all monitoring locations with a similar trend, indicating an issue in the analyses of CrAssphage in this period, even though re-inspection of the procedure and data did not show deviations. Apart from this, differences between flow based, EC based and CrAssphage based normalisation seem to be incidental with a slight underestimation of the EC and CrAss based normalisation, see figure 8. The flow based normalisation results in exactly the same trend as the daily load calculation as applied in the Dutch national sewage surveillance conducted by the National Institute for Public Health and the Environment (SARS-CoV-2 concentration in sample * daily flow per 100.000 inhabitants https://coronadashboard.rijksoverheid.nl/verantwoording#virusdeeltjes-in-rioolwater). The EC based normalisation showed an R^2^ of 0,96 with the daily load, while the CrAss based normalisation results in an R^2^ of 0.92. Flow based normalisation gave the best correspondence with the standard daily load calculation (Castiglioni et al., 2014), while EC based and CrAss based normalisation yielded very comparable results.

**Figure 6.**
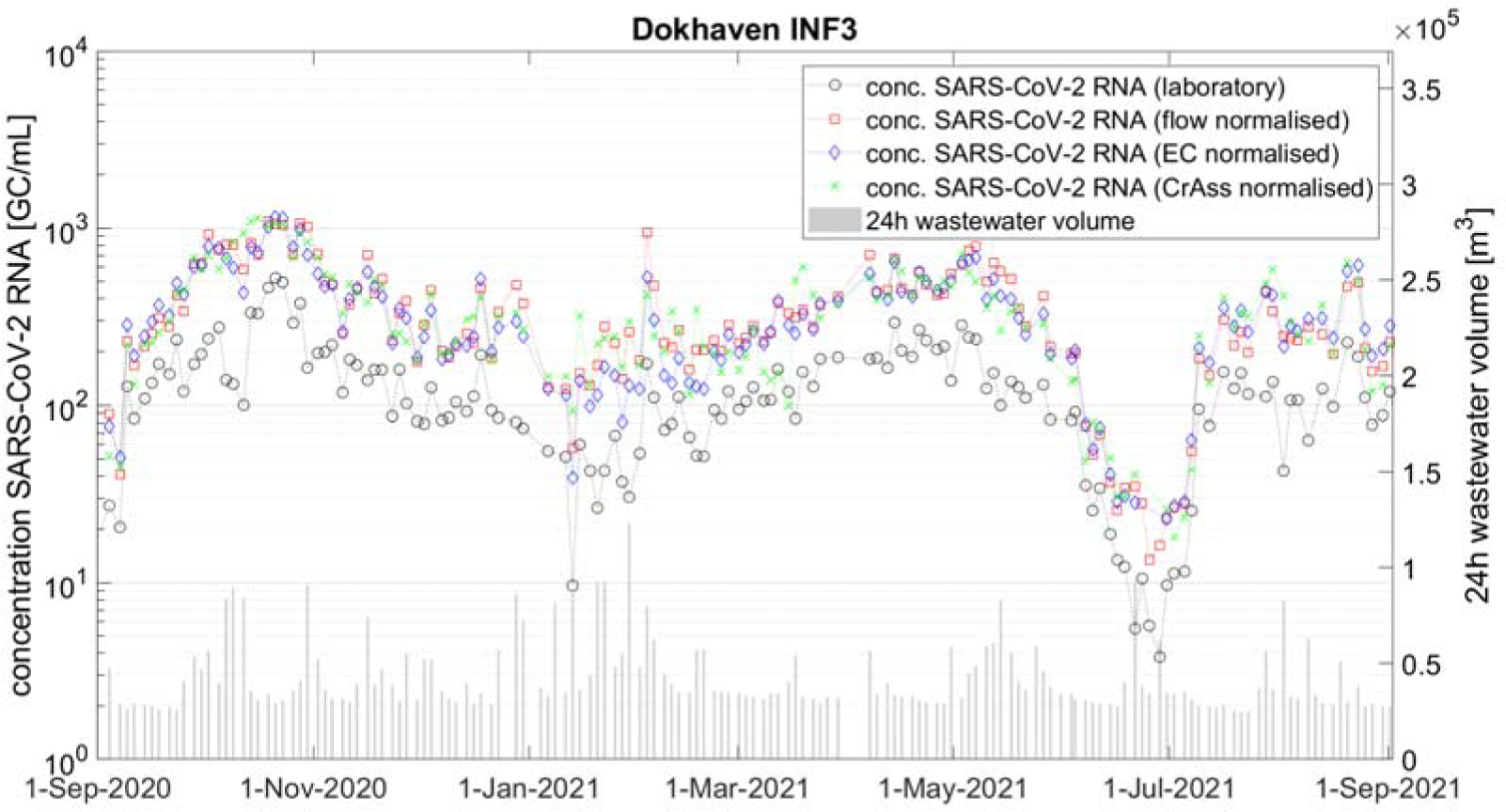
SARS-CoV-2 trends in INF3 catchment. On left axis lab results, flow normalised, CrAss normalised and EC normalised, on right axis the daily flow volume.

**Figure 7.**
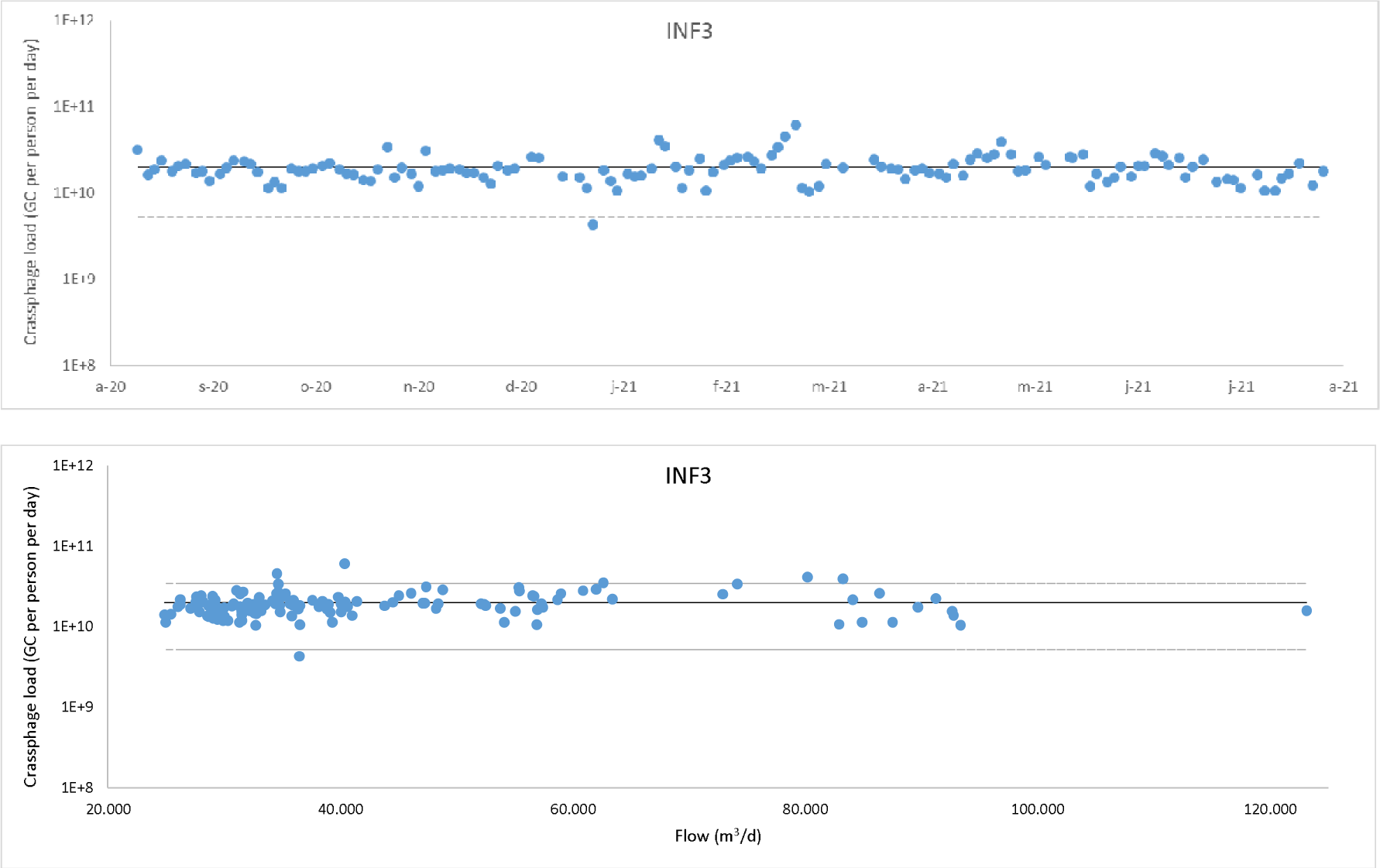
CrAssphage daily load per person in INF3 catchment related to time (top) and related to daily flow (bottom). The solid lines shows the average (2.0 * 10^10^ GC/ml) and the dotted lines are the 2s boundaries (5.2 * 10^9^ – 3.4 * 10^10^ GC/ml).

**Figure 8.**
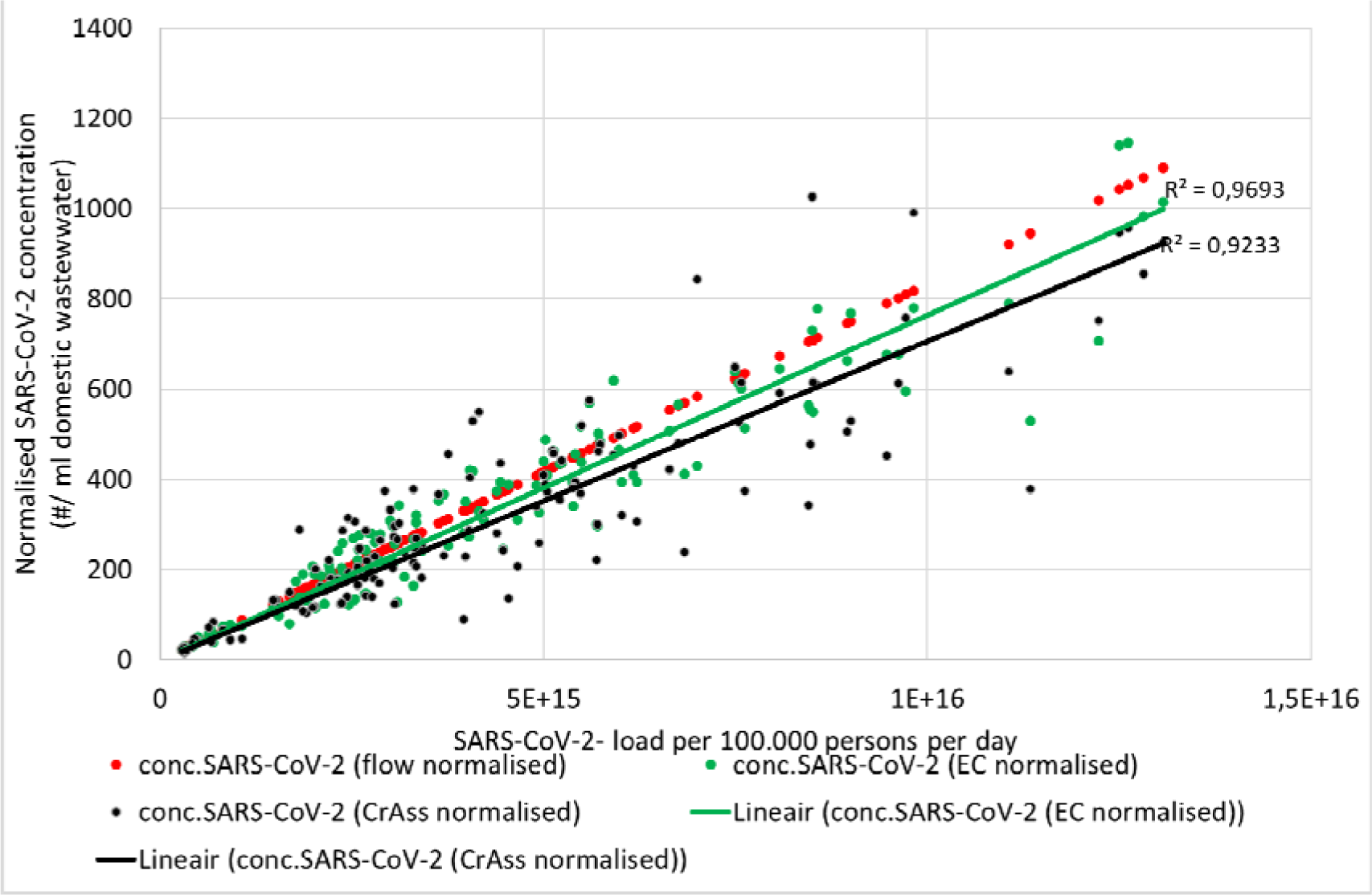
Correlation between daily SARS-CoV-2 load per 100.000 inhabitants and flow based, EC based and CrAss based normalisation for INF3 catchment

### 3.4. Parallel normalisation methods provide quality control

The relative close agreement between flow, EC and CrAssphage based normalisation provides a good basis for quality control of SARS-CoV-2 samples. Of our 1207 SARS-CoV-2 samples, 32 (2.8%) yielded a low CrAssphage concentration (<10^4^ GC/ml). 20 of those were from Ommoord, the site with the most variable proportion of extraneous waters, and these samples also had a high EC (>2000 µS/cm). In these low CrAssphage samples, SARS-CoV-2 N2 concentration was also low or absent. The recovery efficiency of the detection methods for both viruses was low in these samples, possibly due to the presence of humic acids in brackish groundwater coming via clay and peat layers entering the sewer network at this site. Recovery efficiency did not decrease due to high EC due to road de-icing. The data from these samples were therefore excluded from further analysis. Similarly, CrAssphage provided a basis for quality control of detection methods, In August 2020, supply shortages forced us to use other ultrafilters than Centricon. Even though an initial comparison between Centricon and the alternative filters indicated comparable SARS-CoV-2 recoveries, the CrAssphage concentrations obtained in successive wastewater samples processed with these alternative filters were more than 10-fold lower than in the previous sample series. Therefore, data obtained with these alternative filters were not used (data not shown). Moreover, CrAssphage, as a quantitative marker of human feces, also provides the opportunity to check the most basic number in WBE: the population size. When we prepared the monitoring project in summer 2020, we have checked the population size via the municipality of Rotterdam and the Dutch institute of statistics, CBS. These numbers are shown in figure 1. For each monitoring location, we calculated the average Crassphage concentration in dwf samples, shown in SI. The proportion of domestic wastewater of figure 2 agreed to a large extend with the average Crassphage concentration during dwf, except for catchment Katendrecht. A check of the population data of Katendrecht, based on 2020 and 2021 data, learnt that the population in Katendrecht was no longer 4.884 inhabitants, but 5.600 in 2020 and 5.700 in 2021. After adjusting this population number, the adjusted normalisation factors showed good agreement with average Crassphage during dwf.

The EC and CrAssphage data also provided a quality control for the flow normalisation. We developed a procedure to perform the flow and EC based normalisations in parallel. If these two normalisations result in domestic wastewater proportions that differ less than 10%, the flow based normalisation is considered successful and reliable. If the two methods differ more than 10%, it is necessary to analyse the data in more depth and identify the cause of the difference. Figure 9 gives an example for the catchment Katendrecht. Overall, there is a good agreement between the flow and EC based normalisation with a few exceptions. Two of the exceptions occurred close after each other in January 2021. On these days, the pumping station was non-operational between January 19^th^ 14h00 and January 21^st^ 11h00. Consequently, the flow proportional sampling of January 20^th^ only collected samples from 08h00 until the pump failure at 14h00, hence only representing a fraction of the full wastewater production of the area on that day (the other fraction being stored in the sewer system during the pump failure). On January 22^nd^ the opposite occurred: after the re-start, the pumping station processed all wastewater of the preceding 48 hours that had been stored in the sewer system (daily volume: 4635 m^3^), leading to a 24h sample that was not representative of the targeted 24 hours. This is an example of a situation where the flow at the monitoring location was no longer representative of the domestic wastewater produced in the catchment. In both cases, EC based normalisation has been applied instead of flow based normalisation. Figure 9 also shows that on these days, CrAssphage and EC based normalisation would have resulted in similar normalised SARS-CoV-2 concentrations.

**Figure 9.**
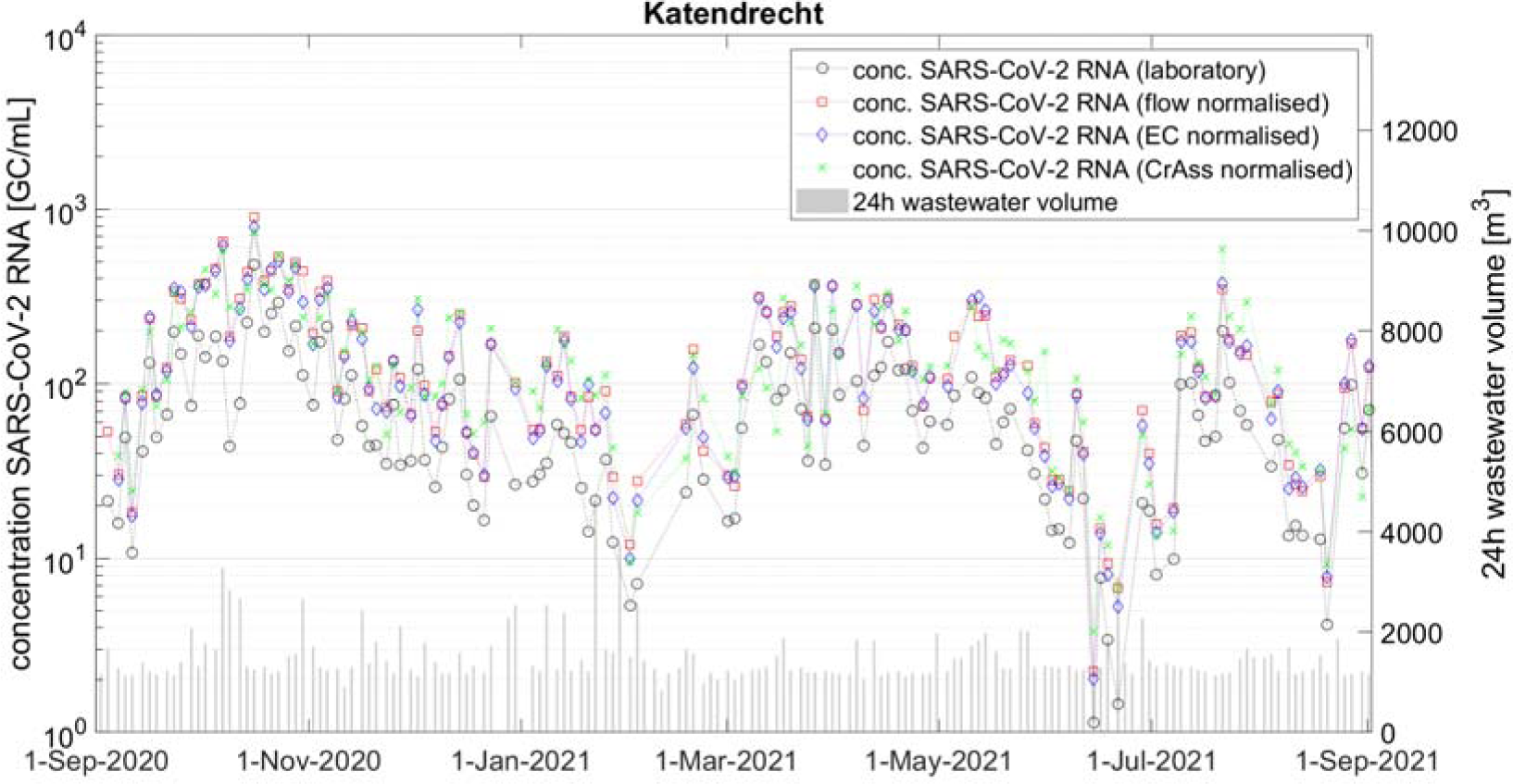
Normalisation of SARS-CoV-2 at the Katendrecht catchment

### 3.5. CrAssphage based normalisation

In order to be able to compare CrAssphage based normalisation with flow and EC based normalisation, flow data were needed to calculate the dwf average value for the CrAssphage and EC normalisation. In situations where flow data is unavailable, it is not possible to first derive this dwf average to serve as a reference for calculation dilution of the domestic wastewater with industrial wastewater, runoff and extraneous waters. In those situations, direct normalisation using the ratio SARS-CoV-2/CrAssphage can be applied, as CrAssphage based normalisation using the dwf average renders the exact same SARS-CoV-2 concentration trends as using the ratio SARS-CoV-2/CrAssphage. In the catchments of Rotterdam, with population varying from 5.600 to 138.280, the CrAssphage load per person per day was very comparable between city areas (Figure 10) and constant over time (Figure 7 and Figures S2), making CrAssphage suitable for direct normalisation for the human faecal fraction in wastewater samples at the population sizes in this study (5.600 and up).

**Figure 10.**
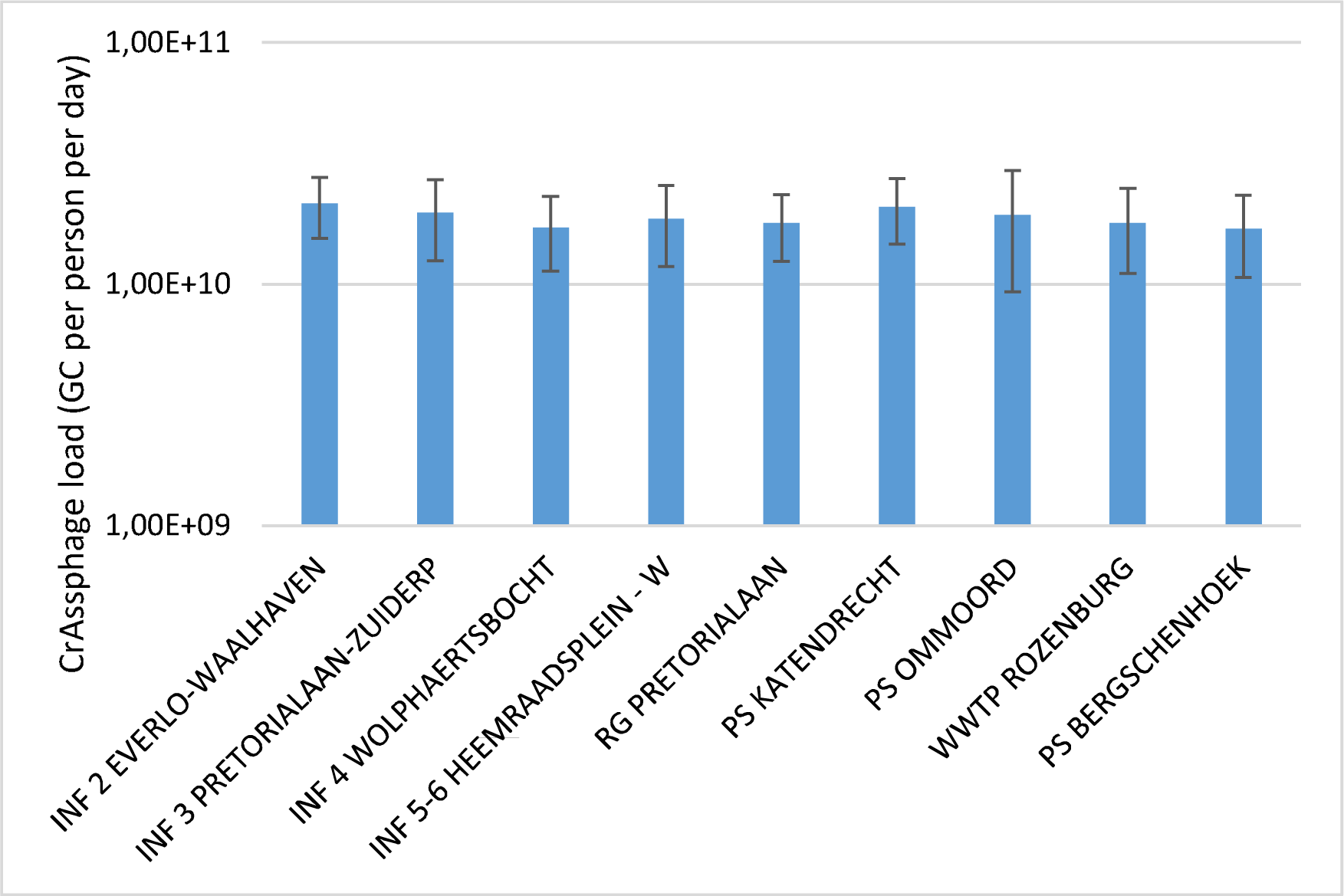
Average CrAssphage load per person per day in the different city areas. Error bars show the standard deviation.

### 3.6. CrAssphage in stool samples

Stool samples were collected from SARS-CoV-2 positive adults (N=221) that notified their general practitioner of a positive SARS-CoV-2 test performed at the GGD testing streets. The CrAssphage concentrations in these stool samples showed very large variation, spanning more than 10 log^10^ units (Figure 11). A small percentage (4.5%) had no detectable (<140 GC/ml faeces) CrAssphage in their stool sample. In the positive stool samples, a bimodal distribution was observed, with peaks at 10^3.5^ and at 10^8.5^ GC/ml faeces.

**Figure 11.**
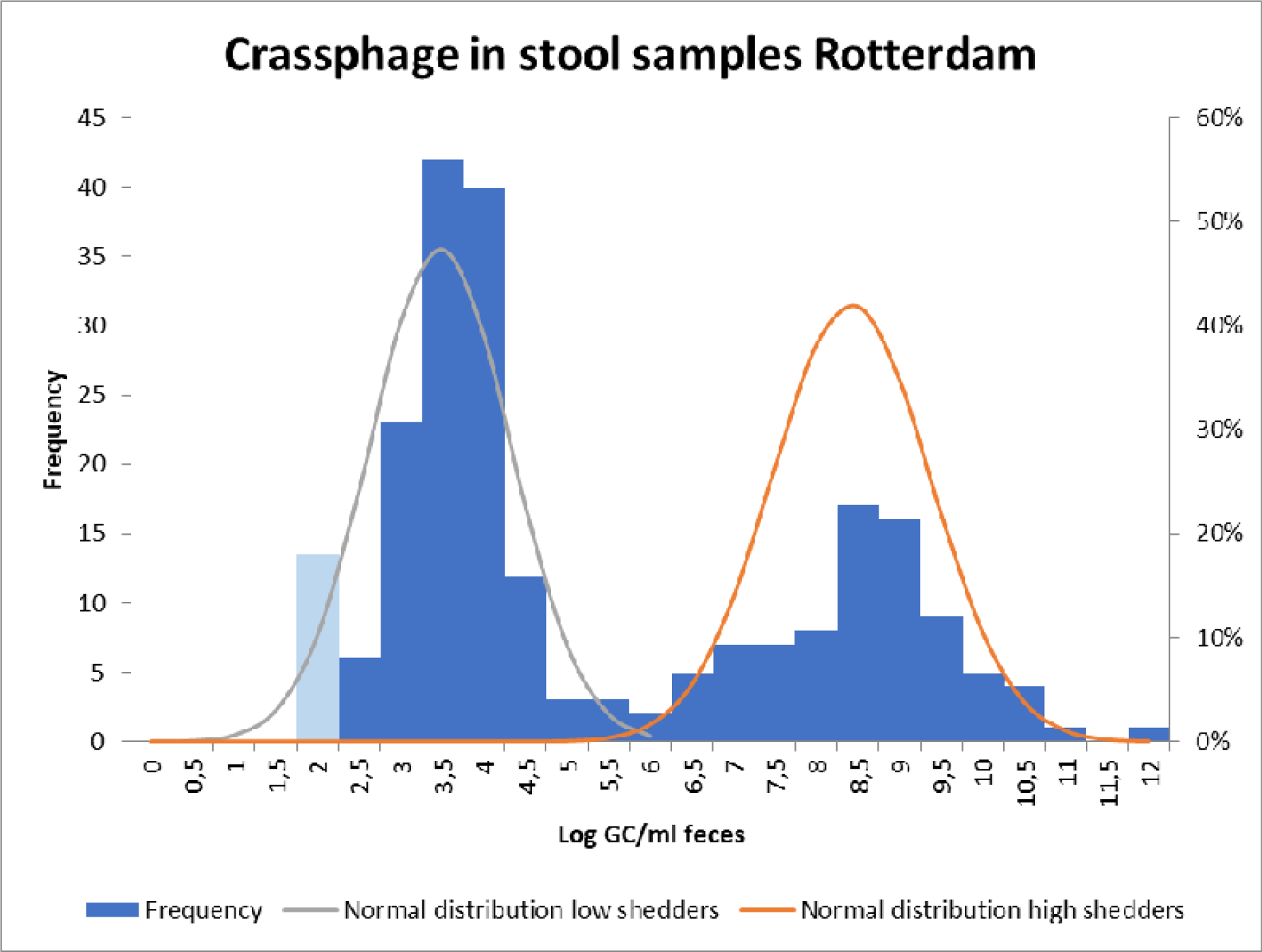
Distribution of CrAssphage GC/ml in human stool samples (n=221) from Rotterdam. Frequency of data below the detection limit (140 GC/ml) is shown in light blue. Normal distribution fitted through the log-transformed concentrations in stool samples of the low shedder (<10^6.5^ GC/ml; mean: 3.474; standard deviation 0.843; in grey) and the high shedder (≥10^6.5^ GC/ml; mean: 8.413; standard deviation 0.950; in orange).

The arithmetic average concentration of CrAssphage in stool samples is 3.39 x 10^9^ GC/ml faeces. This average was largely determined by a single stool sample contained an extremely high concentration (10^12^/ml). The high concentration was confirmed by re-analysis of the sample. Because of the bimodal distribution of CrAssphage in stool samples, the very high concentration in the small proportion of samples is dominant in determining the arithmetic average. To be less dependent on data from a single stool sample, the shedding data were fitted to two separate Normal distribution after log-transformation, one for the low shedding part of the population (66.1%, including the non-detects), with an average of 3.474 LOG GC/ml and a standard deviation of 0.843 LOG GC/ml, and one for the high shedding part (33.9%, with an average of 8.413 LOG GC/ml and a standard deviation of 0.950 LOG GC/ml. With the average of the high shedding proportion of the population, and an average stool production of 128 g wet weight (≈ml) pppd (Rose et al., 2015), the average CrAssphage load of the population would be 1.12 x 10^10^ GC pppd. This is close to the average measured CrAssphage load in the wastewater of the city areas of 1.9 * 10^10^ pppd. CrAssphage is quite stable in the environment (Balleste et al, 2019). The lack of correlation between daily flow and daily CrAssphage-load per person (Figure 7) suggests that there is no significant build-up of CrAssphage in sediments or biofilm in the sewer network (Balleste et al., 2019), unlike for other parameters such as COD and TSS (Schilperoort et al., 2012), see also figures in S3, or pollutants that show a strong adsorption to TSS (Gasperi et al., 2014).

Compared to the variability of CrAssphage shedding per person, the observed CrAssphage concentration in wastewater was very comparable between different city areas and over time. This indicates that, even though the variation in CrAssphage shedding between individuals is very high, the ‘population shedding’ is fairly constant in population sizes of 5.600 and above. Although CrAssphage has been shown to reside in the human gut for months (Siranosian et al., 2020; Honap et al., 2020; Tamburini et al., 2021), little is known about the CrAssphage shedding dynamics per person over time. A longitudinal study in three infants showed considerable variation over the first year of life (Taboada et al, 2021), but a study among ten healthy adults showed that the human gut virome composition is personalized and relatively stable over time, including for CrAssphage (Shkoporov et al., 2019). The high variability in CrAssphage concentration in stool samples observed in this study, would imply that CrAssphage would become less suitable as normaliser in small populations.

## 4. Conclusions

This study showed that without normalisation SARS-CoV-2 sewage data may misrepresent the actual short-term trends of COVID-19 circulation in the population due to the impact of rain and snowmelt in a combined sewer network. Unnormalised data do show similar long-term trends as normalized data, but it is the short-term (1-2 weeks) trends are the most relevant to support public health actions to limit the transmission of the virus and where discrepancies were considerable. Also, for comparison of SARS-CoV-2 circulation in city areas (or cities), normalisation is needed to compensate for the different proportions of extraneous water in different (parts of the) sewer networks.

Flow appears to be most appropriate normalizer, for only a few examples in our many data (2/1207; 0.17%) flow was not appropriate, due to sewer blocking. Since EC captured these errors, we propose flow-based normalisation in combination with an EC check. In our study, EC had the added value of picking up brackish groundwater intrusion and road de-icing, the latter being associated with reduced recoveries of the CrAssphage and SARS-CoV-2 detection methods.

The CrAssphage loads per population per day was very comparable between city areas and constant over time, making CrAssphage-based normalisation an adequate alternative normalization procedure in the absence of flow data or in situations where flow data become unreliable. CrAssphage could also be used to normalize the results of passive samplers, an upcoming method of monitoring SARS-CoV-2 (Shang et al., 2021) by serving as index for the human fecal matter captured in the samplers. CrAssphage shedding per person in stool data was highly variable. This would imply that CrAssphage normalization may become unreliable if the sampled population is small. Further research on the limits of application is necessary.

## Data Availability

All data produced in the present work are contained in the figures of the manuscript

## Acknowledgements

The authors are very grateful for the assistance of the Water Authorities and WWTP and pumping station operators of Waterschap Hollandse Delta and municipality of Rotterdam, staff of IMD for installing and maintaining the autosamplers and sampling specialists of AQUON, Toke Mulder – van Kempen and Eline Hoogteijling for assisting with the inclusion of the patients for the fecal sampling study. This study was financed by STOWA, TKI Watertechnology in collaboration with Erasmus Foundation, Adessium Foundation, European Union’s Horizon H2020 grant VEO (grant no. 874735), Ministry of Health, Welfare and Sport, H2020 and Waterboards Waterschap Hollandse Delta, Hoogheemraadschap van Delfland and Hoogheemraadschap Schieland en Krimpenerwaard. The authors would like to thank all the patients that contributed to the fecal sampling study as well as all the included GP-practices.

