## supplemental figures for "Normalisation of SARS-CoV-2 concentrations in wastewater: the use of flow, conductivity and CrAssphage"

### Supplemental material

#### S1. Normalisation results of SARS-CoV-2 concentration per catchment

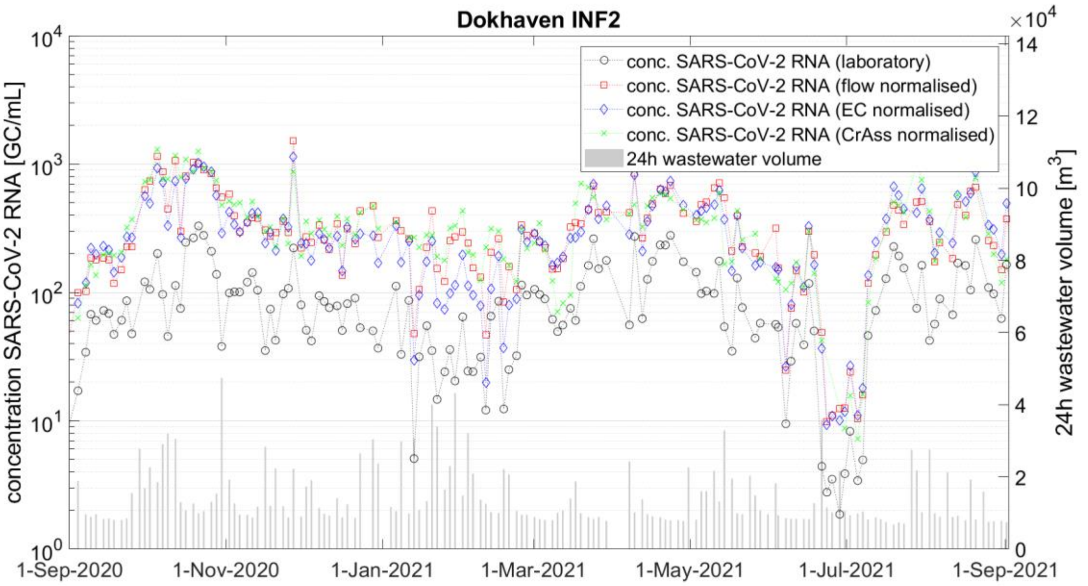


Figure S1.1. SARS-CoV-2 trends in INF2 catchment. On left axis lab results, flow normalised, CrAss normalised and EC normalised, on right axis the daily flow volume.

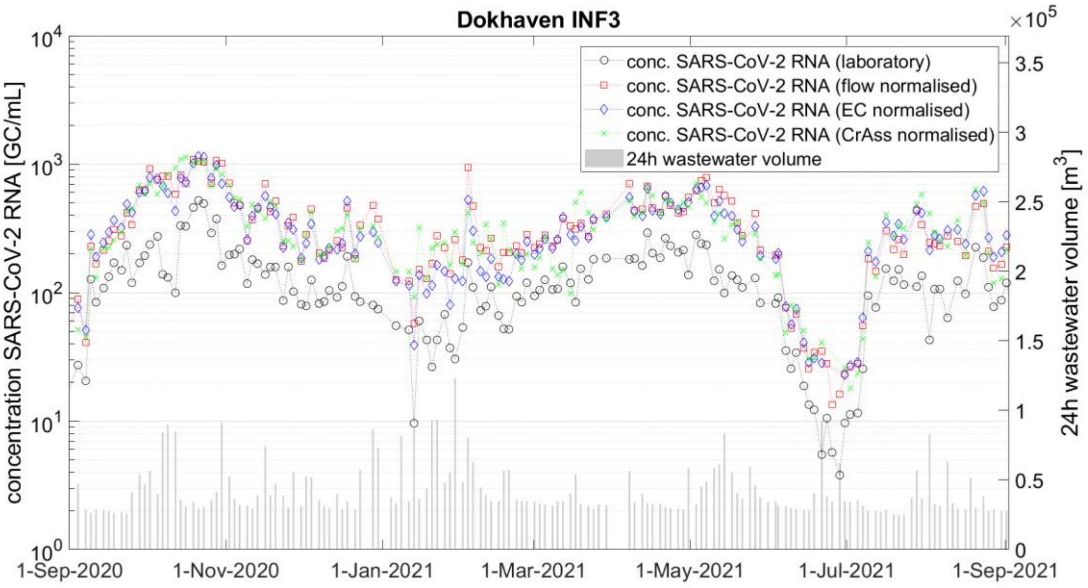

Figure S1.2. SARS-CoV-2 trends in INF3 catchment. On left axis lab results, flow normalised, CrAss normalised and EC normalised, on right axis the daily flow volume.

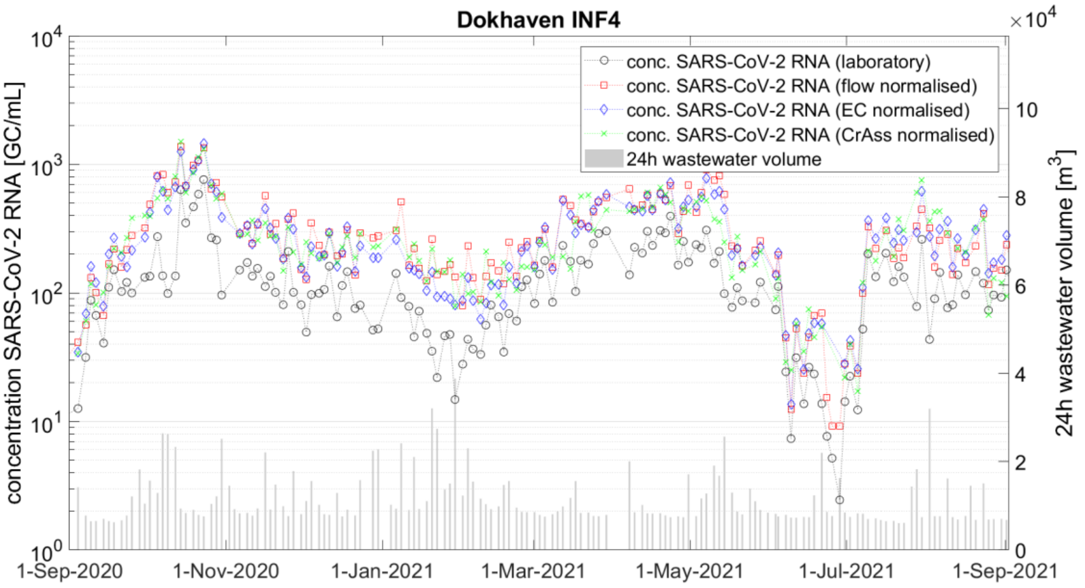


Figure S1.3. SARS-CoV-2 trends in INF4 catchment. On left axis lab results, flow normalised, CrAss normalised and EC normalised, on right axis the daily flow volume.

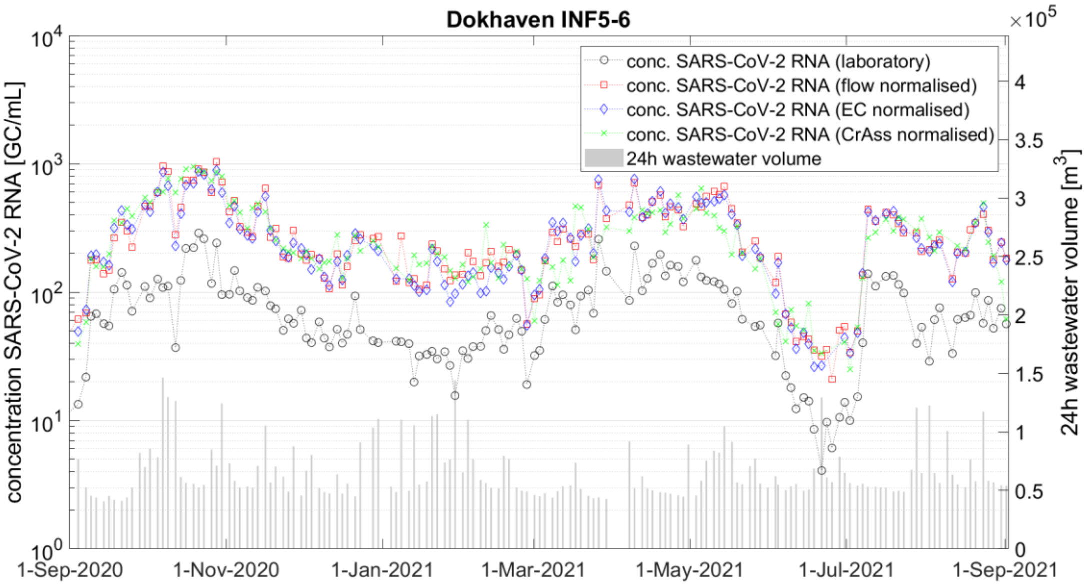

Figure S1.4. SARS-CoV-2 trends in INF5-6 catchment. On left axis lab results, flow normalised, CrAss normalised and EC normalised, on right axis the daily flow volume.

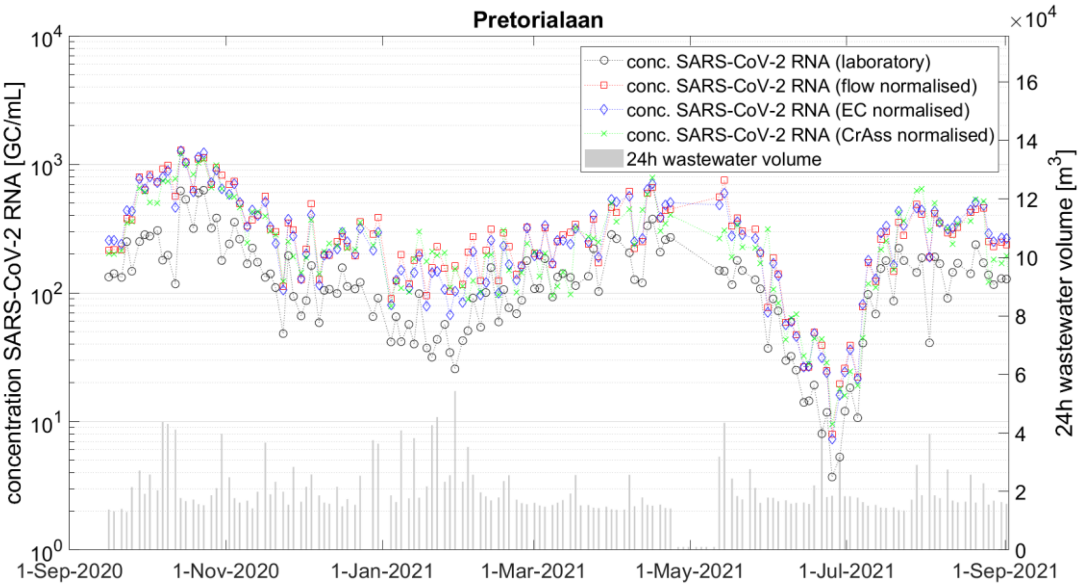


Figure S1.5. SARS-CoV-2 trends in Pretorialaan catchment. On left axis lab results, flow normalised, CrAss normalised and EC normalised, on right axis the daily flow volume.

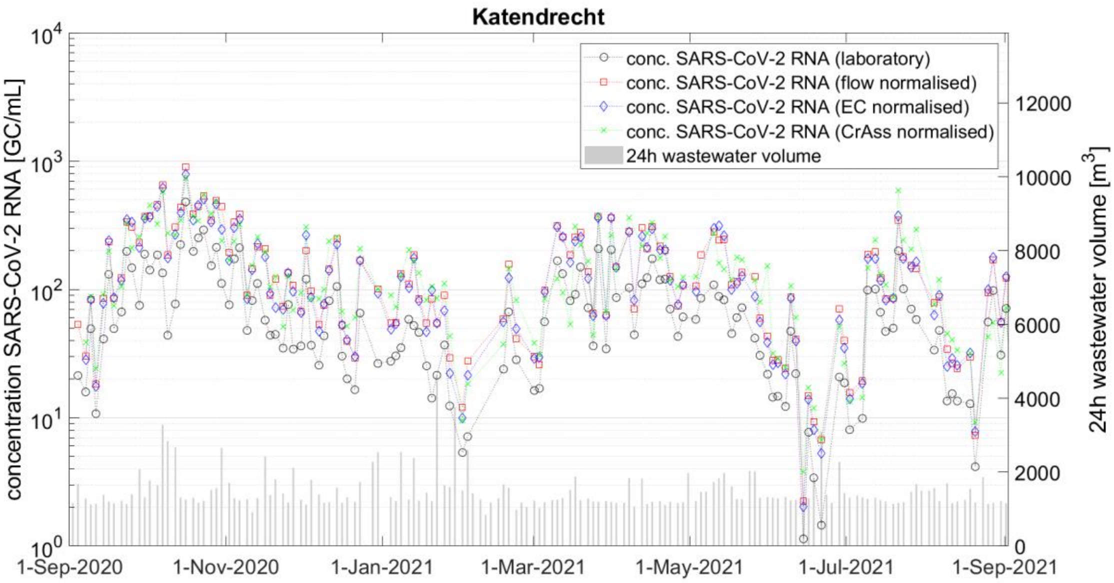

Figure S1.6. SARS-CoV-2 trends in Katendrecht catchment. On left axis lab results, flow normalised, CrAss normalised and EC normalised, on right axis the daily flow volume.

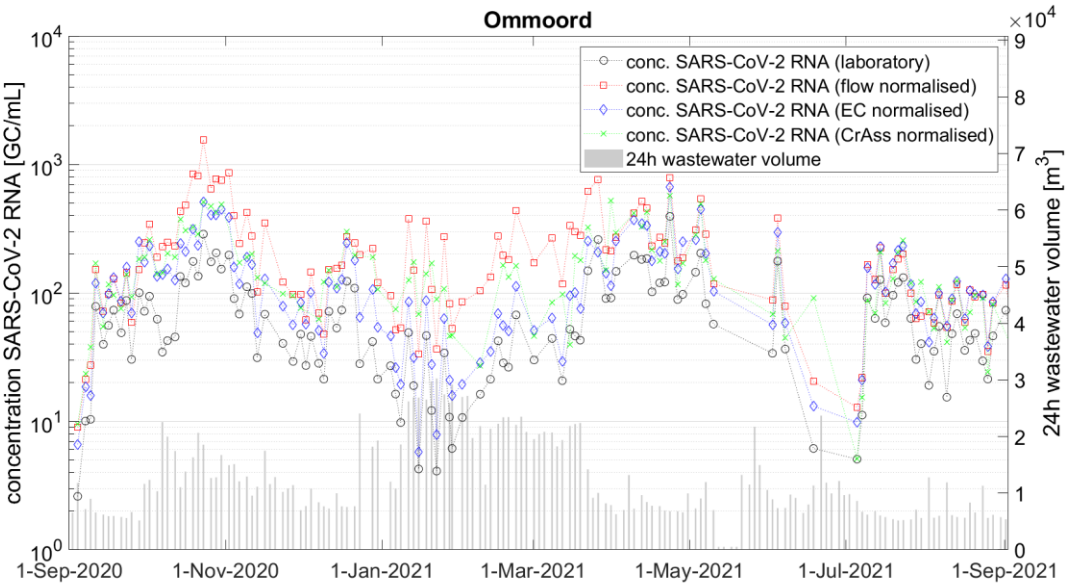


Figure S1.7. SARS-CoV-2 trends in Ommoord catchment. On left axis lab results, flow normalised, CrAss normalised and EC normalised, on right axis the daily flow volume.

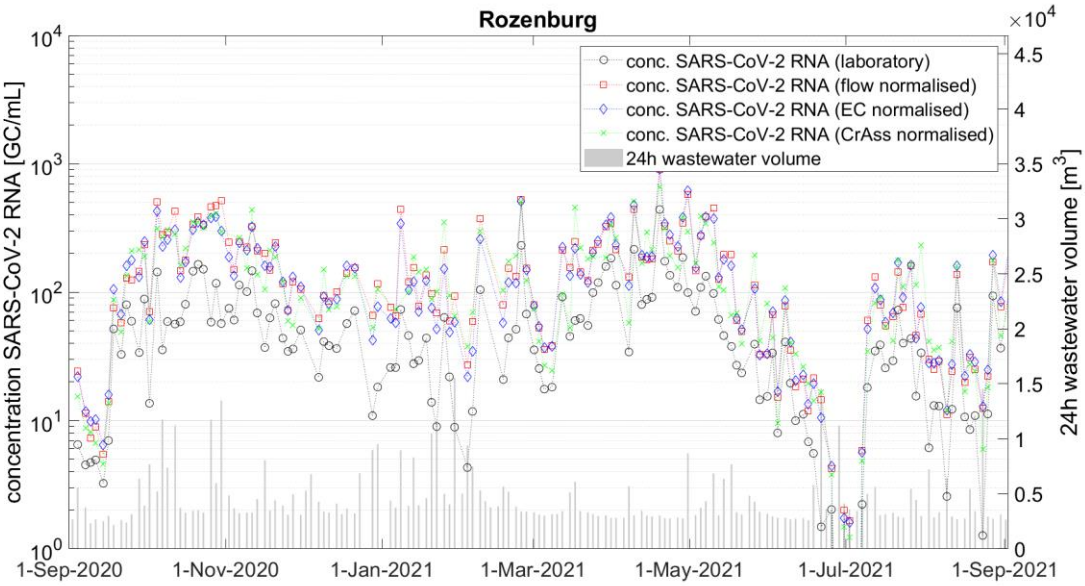

Figure S1.8. SARS-CoV-2 trends in Rozenburg catchment. On left axis lab results, flow normalised, CrAss normalised and EC normalised, on right axis the daily flow volume.

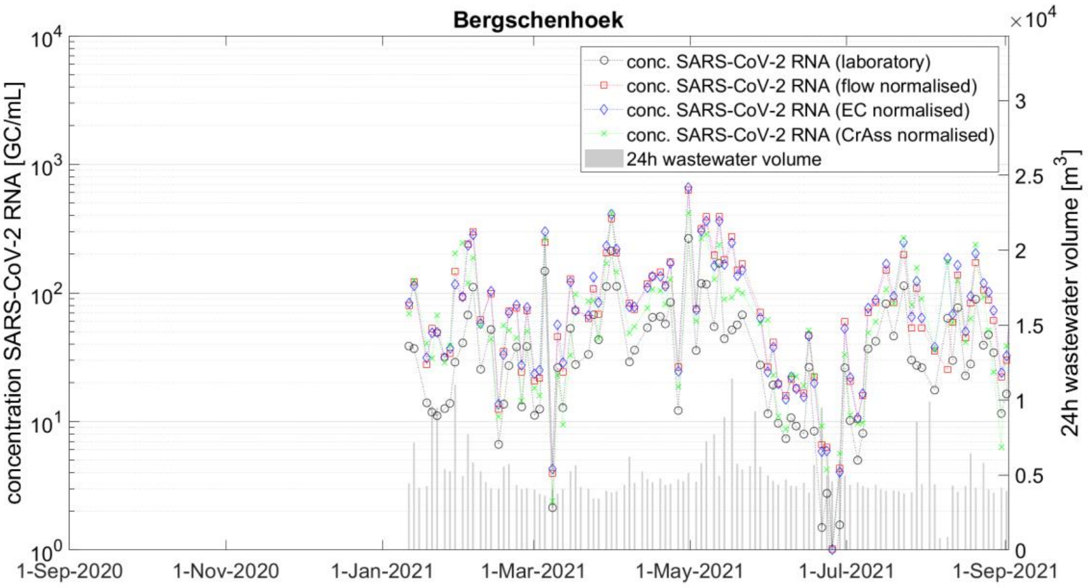


Figure S1.9. SARS-CoV-2 trends in Bergschenhoek catchment. On left axis lab results, flow normalised, CrAss normalised and EC normalised, on right axis the daily flow volume.

#### S2. Crassphage daily load per person for each catchment

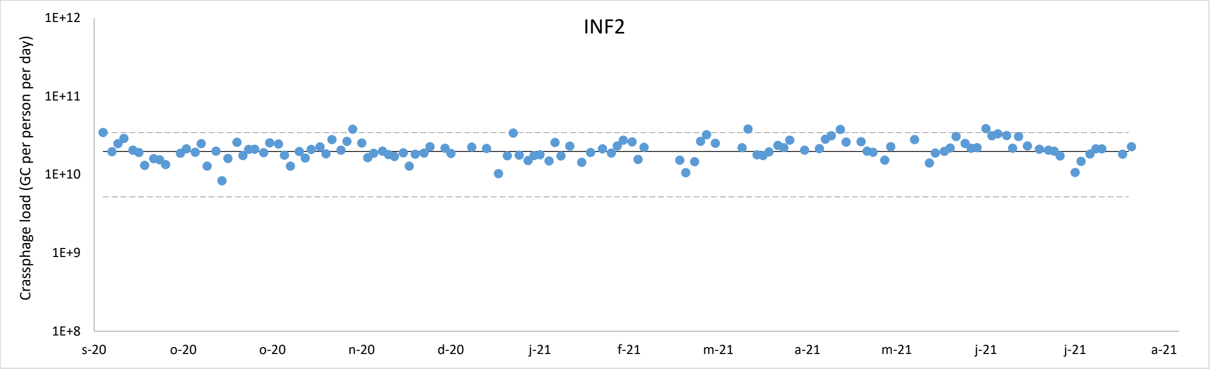


Figure S2.1. CrAssphage daily load per person in INF2 catchment. The solid line shows the average and the dotted lines are the 2s boundaries.

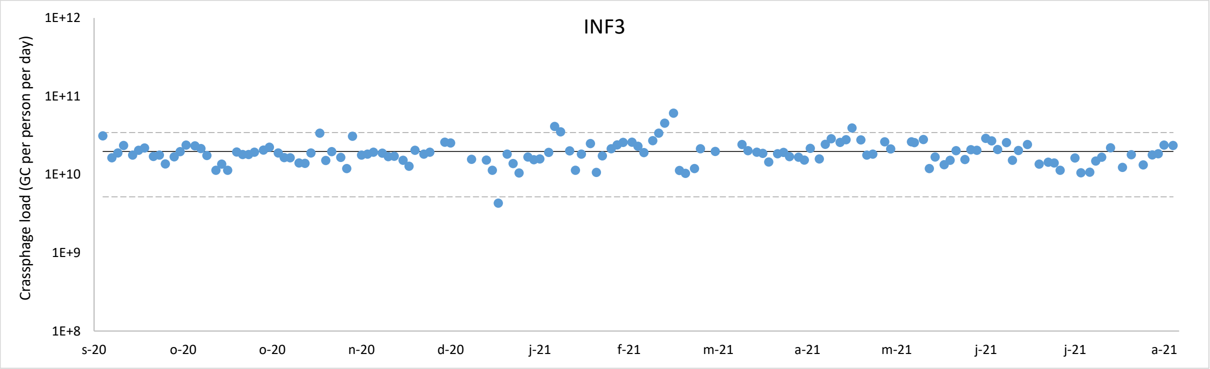


Figure S2.2. CrAssphage daily load per person in INF3 catchment. The solid line shows the average and the dotted lines are the 2s boundaries

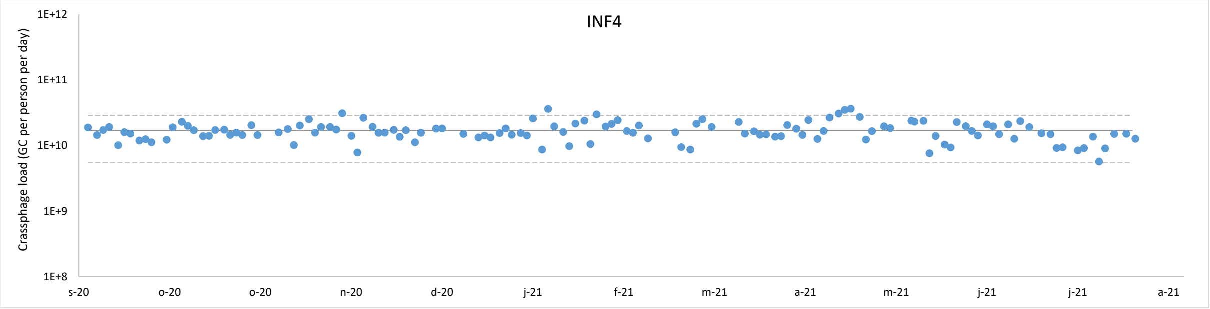


Figure S2.3. CrAssphage daily load per person in INF4 catchment. The solid line shows the average and the dotted lines are the 2s boundaries

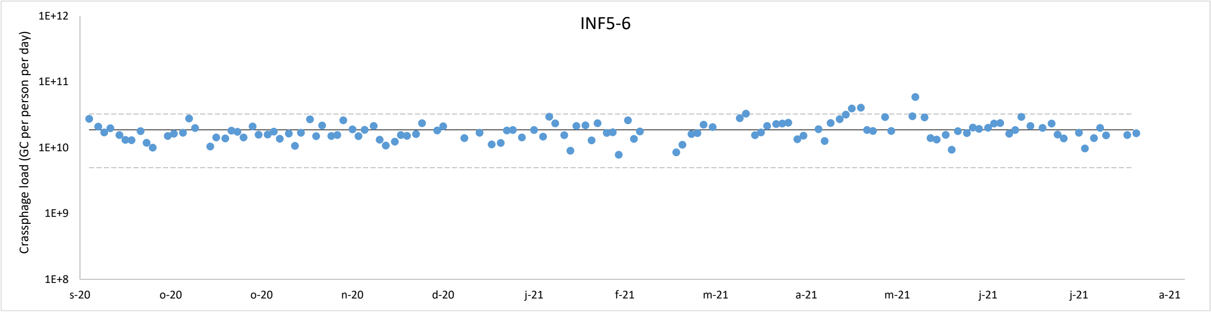


Figure S2.4. CrAssphage daily load per person in INF5-6 catchment. The solid line shows the average and the dotted lines are the 2s boundaries

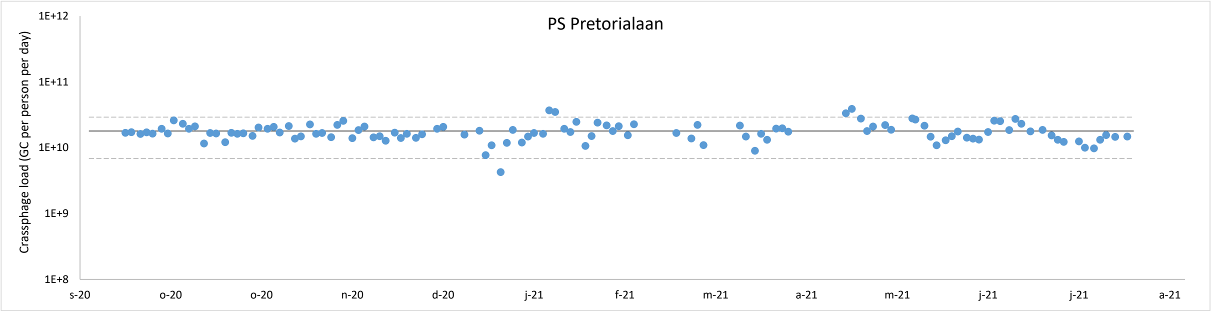


Figure S2.5. CrAssphage daily load per person in PS Pretorialaan catchment. The solid line shows the average  and the dotted lines are the 2s boundaries

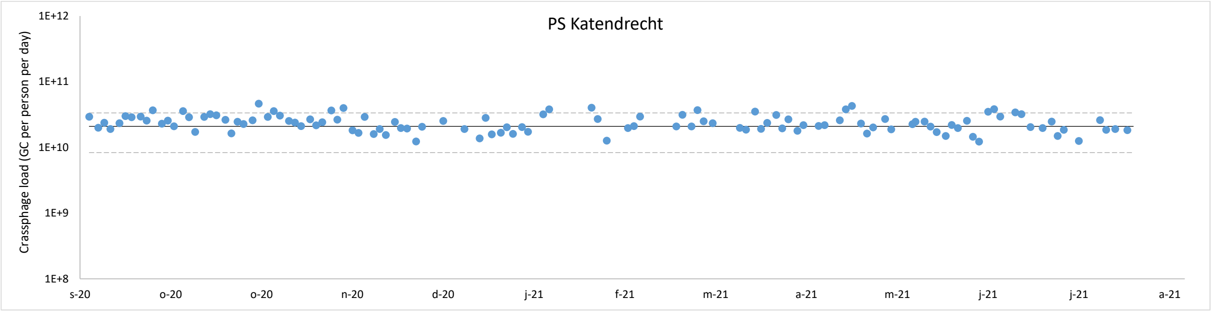


Figure S2.6. CrAssphage daily load per person in PS Katendrecht catchment. The solid line shows the average  and the dotted lines are the 2s boundaries

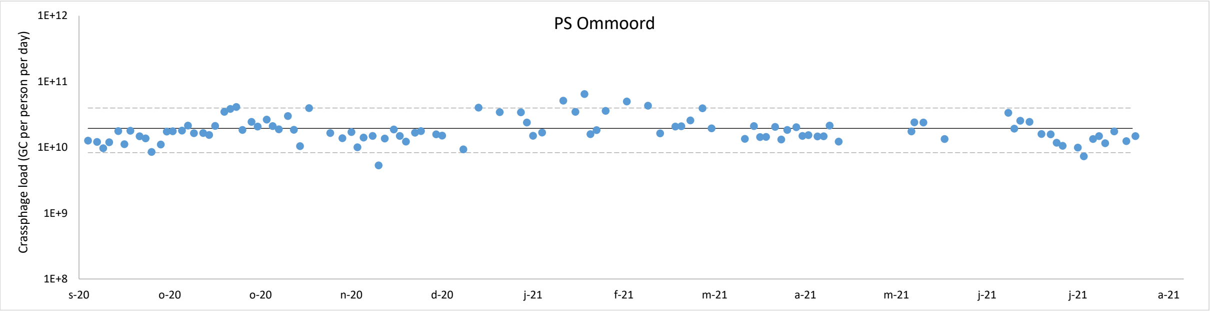


Figure S2.7. CrAssphage daily load per person in Ps Ommoord catchment. The solid line shows the average and the dotted lines are the 2s boundaries

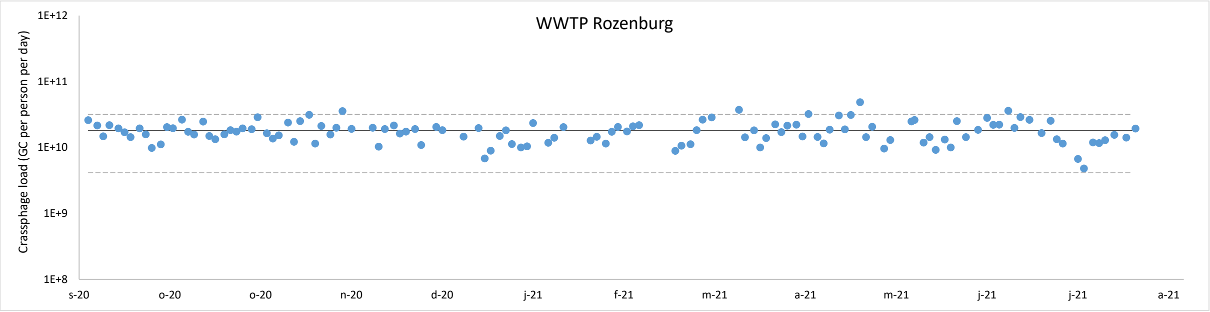


Figure S2.8. CrAssphage daily load per person in WWTP Rozenburg catchment. The solid line shows the average and the dotted lines are the 2s boundaries

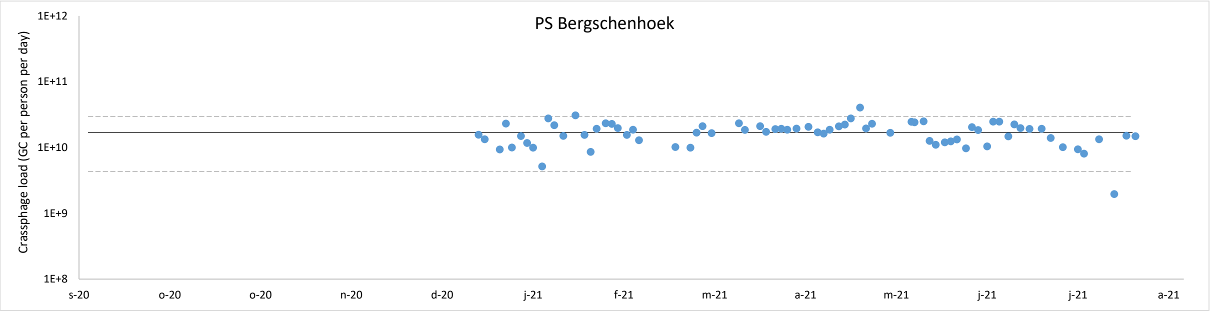


Figure S2.9. CrAssphage daily load per person in PS Bergschenhoek catchment. The solid line shows the average and the dotted lines are the 2s boundaries

#### S3. Daily loads of standard wastewater parameters versus daily flows

The figures in S3 show the relation between daily flow and traditional wastewater parameters for the influent of wwtp Dokhaven. The load during WWF is higher for all parameters due to in sewer stocks and, to a minor extent, storm water runoff.

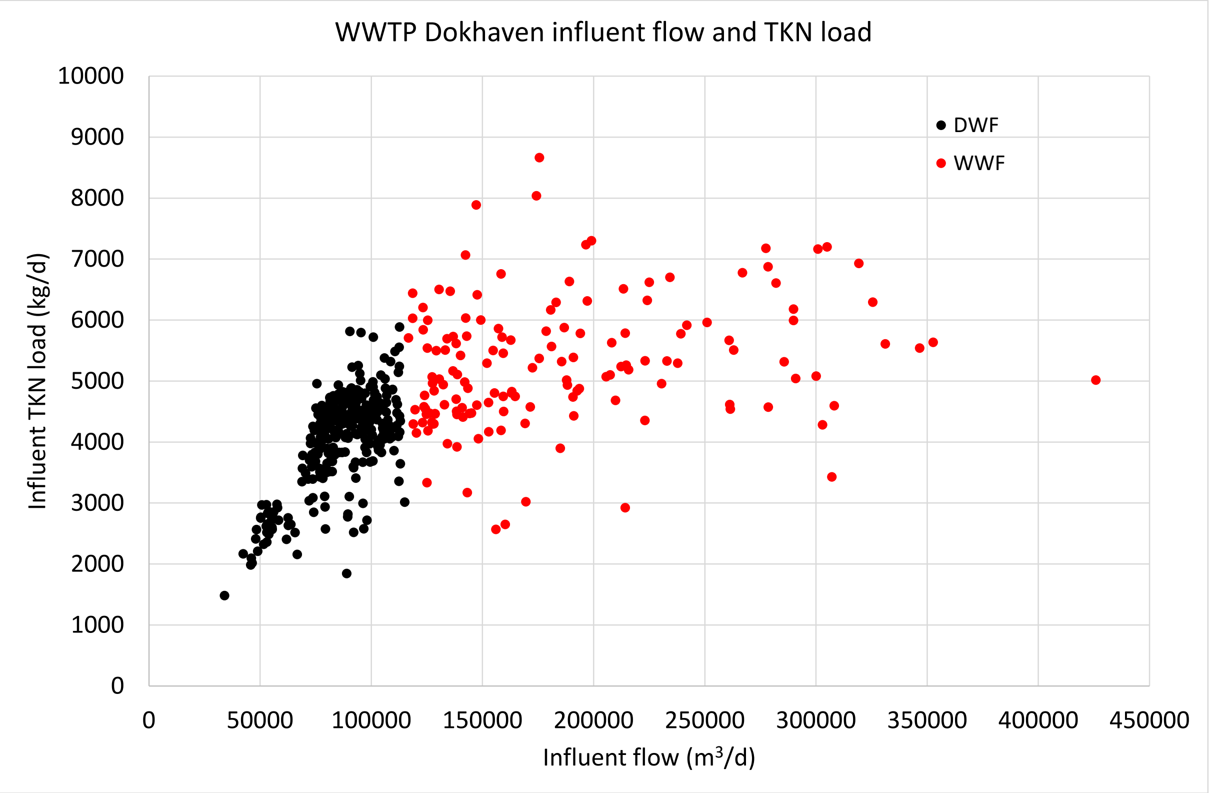


Figure S3.1. TKN load in influent wwtp Dokhaven. WWF average = 1.3*DWF average

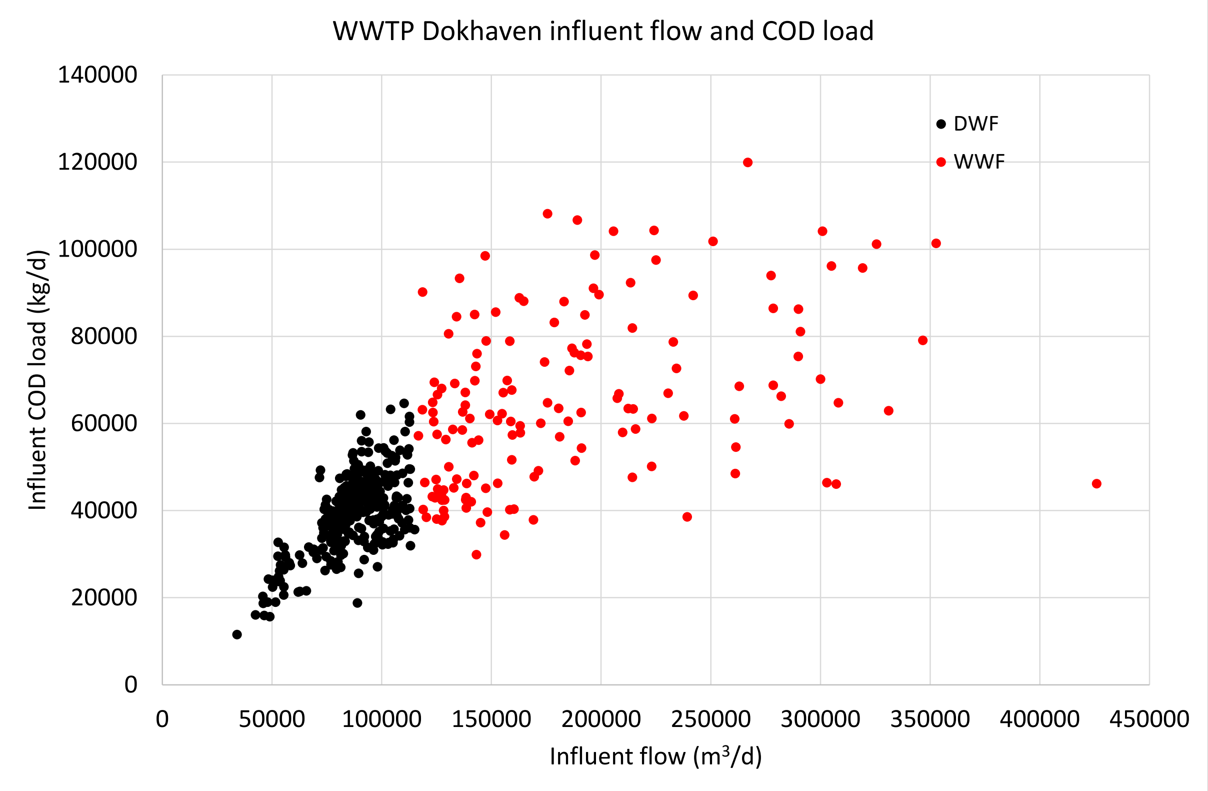


Figure S3.2. COD load in influent wwtp Dokhaven. WWF average = 1.7*DWF average

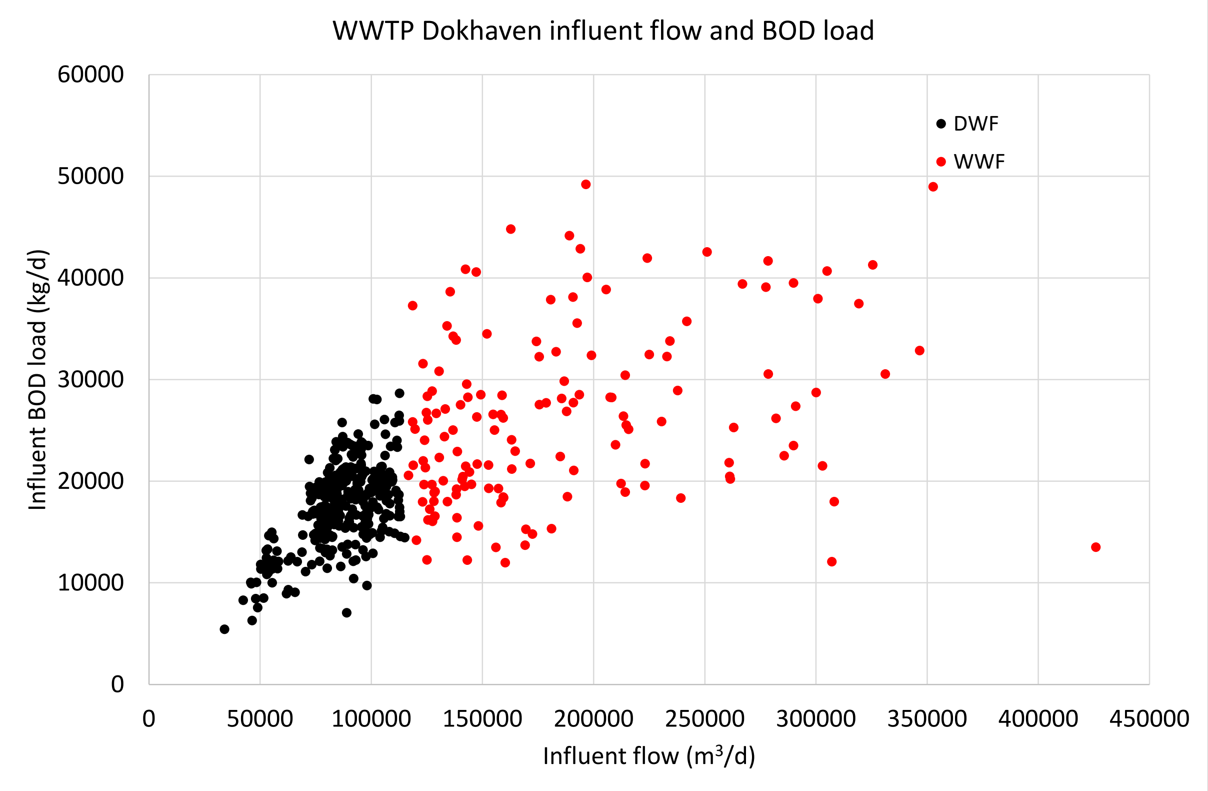


Figure S3.3. BOD load in influent wwtp Dokhaven. WWF average = 1.5*DWF average

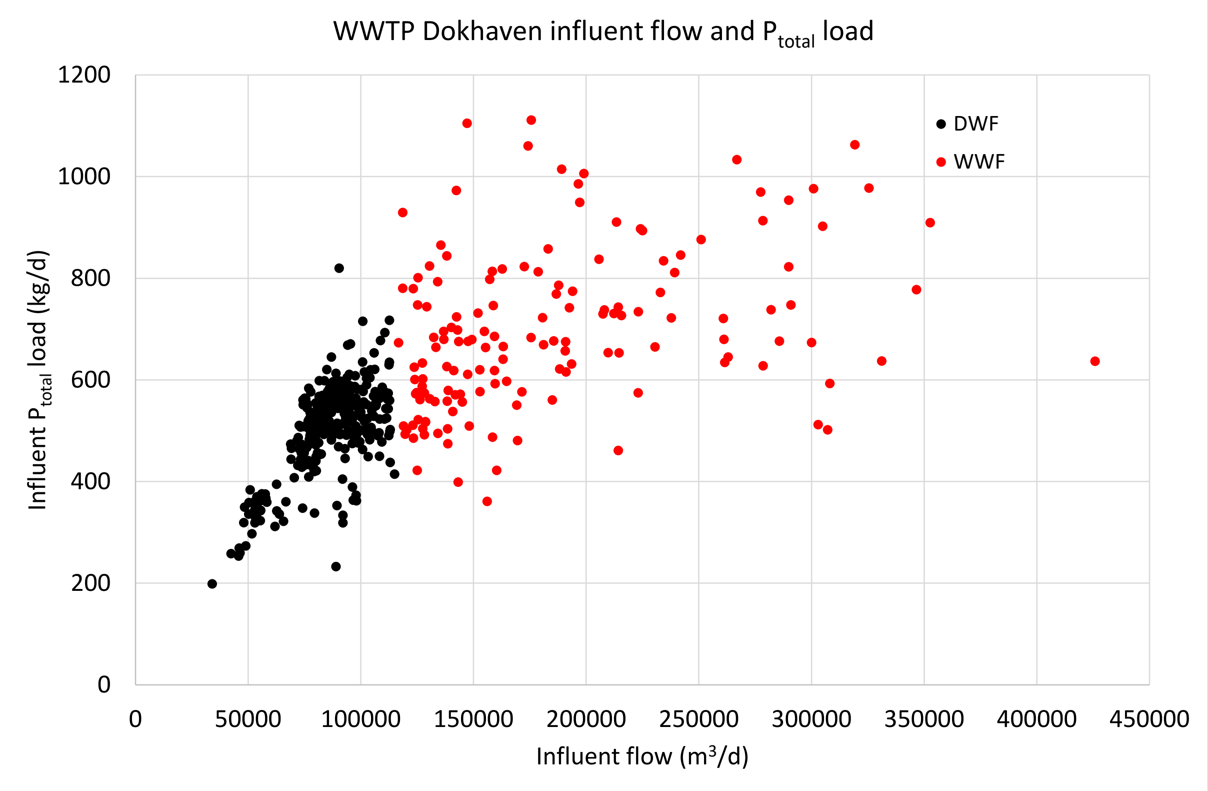


Figure S3.4. $P_{total}$ load in influent wwtp Dokhaven. WWF average = 1.4*DWF average

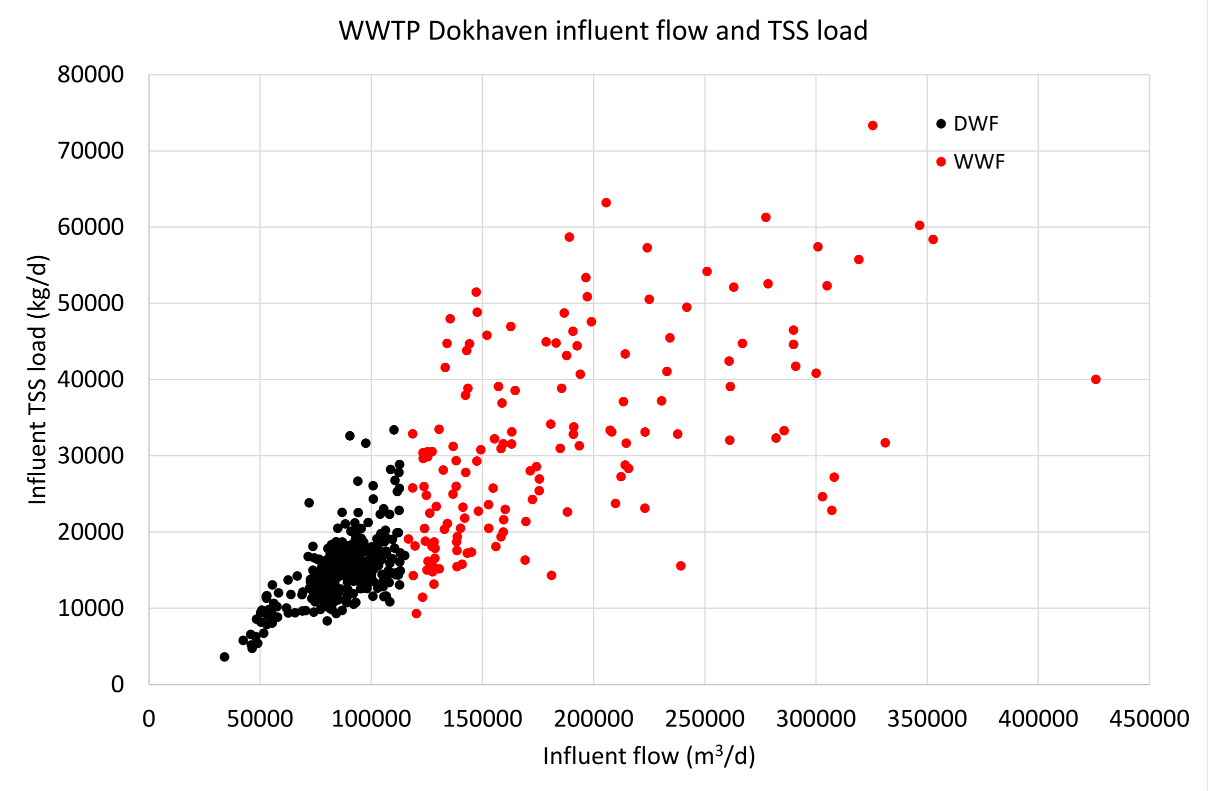


Figure S3.5. TSS load in influent wwtp Dokhaven. WWF average = 2.2*DWF average
